# Socially stratified DNA-methylation profiles are associated with disparities in child and adolescent mental health

**DOI:** 10.1101/2021.09.17.21263582

**Authors:** L. Raffington, P. Tanksley, L. Vinnik, A. Sabhlok, M.W. Patterson, T. Mallard, M. Malanchini, Z. Ayorech, E.M. Tucker-Drob, K.P. Harden

**Author notes:** Corresponding Authors: Laurel Raffington, University of Texas at Austin, 108 E. Dean Keeton Stop A8000, Austin, Texas 78712-1043., Kathryn P. Harden, University of Texas at Austin, 108 E. Dean Keeton Stop A8000, Austin, Texas 78712-1043. **These authors contributed equally**: Elliot M. Tucker-Drob, Kathryn P. Harden.

## Abstract

**Importance:** Economic and racial inequality is linked to disparities in children’s mental health. Biomarkers that reflect these social disparities are lacking.

**Objective:** We examined the hypothesis that salivary DNA-methylation patterns of higher inflammation and faster pace of biological aging are economically, racially and ethnically stratified and are associated with child mental health.

**Design:** The Texas Twin Project is an on-going, observational, longitudinal study that began in May 2012. Analyses were preregistered on May 7, 2021, and completed on August 23, 2021.

**Setting:** The population-based study identified and recruited participants from public school rosters in the greater Austin area.

**Participants:** Participants in the analytic data set included all participants that agreed to contribute DNA samples and whose samples were assayed by January 2021.

**Exposures:** Family- and neighborhood-level socioeconomic inequality, racial and ethnic identities (White, Latinx, Black, Asian).

**Main Measure(s):** Environmental exposures were analyzed in relation to salivary DNA-methylation profiles of higher inflammation (DNAm-CRP) and faster pace of biological aging (DunedinPoAm). Child internalizing problems, attention problems, aggression, rule-breaking, ADHD, oppositional defiant disorder, and conduct disorder were measured using parent-reports and self-reports on abbreviated versions of the Achenbach Child Behavior Checklist and Conners 3.The hypotheses being tested were formulated after data collection of the present data freeze and were pre-registered prior to analyses being conducted.

**Results:** In a sample of *N*=1,183 8-to-19-year-olds (609 female, age M=13.38y), children’s salivary DNA-methylation profiles and psychiatric symptoms differed by socioeconomic conditions, race and ethnicity. Children with more parent-reported internalizing symptoms had higher DNAm-CRP (*r*=0.15, 95% CI=0.05 to 0.25, P=0.004) and DunedinPoAm (*r*=0.15, CI=0.05 to 0.25, P=0.002), and children with more parent-reported aggression problems had higher DNAm-CRP (*r*=0.17, CI=0.04 to 0.31, P=0.013). DNAm-CRP partially mediated advantage of higher family socioeconomic status (16% of total effect) and White racial identity (12% of total effect) on reduced internalizing symptoms. DunedinPoAm also partially mediated advantage of White racial identity on internalizing (19% of total effect).

**Conclusions and Relevance:** Socioeconomic and racial inequality are visible in children’s epigenetic profiles of inflammation and the rate of biological aging in a manner that is tied to social disparities in mental health.

**Key Points:** *Question:* We examined whether salivary DNA-methylation profiles are socially stratified and associated with child mental health.

*Findings:* In this preregistered, cross-sectional observational study of 1,183 children and adolescents, socioeconomic, racial, and ethnic disparities in mental health were associated with salivary DNA-methylation profiles of inflammation and the pace of biological aging.

*Meaning:* DNA-methylation biomarkers hold promise as tools to quantify the biological impact of socioeconomic inequality and being racially minoritized in a manner that is tied to social disparities in mental health.

## 3. Text

Children who experience environmental adversities, including financial scarcity and the discrimination, prejudice, and oppression associated with racial and ethnic marginalization, are at increased risk of experiencing symptoms of psychiatric disorders. Their mental health burden includes both problems with internalizing such as anxiety and depression, as well as behaviors characteristic of externalizing disorders like aggression and hyperactivity^1–3^. In contrast, White racial identity may confer protective effects on children’s mental health, as White children experience the generational legacy of state-sanctioned social power, resources, and favoritism lived by White people, *i.e.* White privilege^4, 5^. We capitalize these terms to highlight that racial and ethnic identities are social constructions that are not based on “innate” biosocial boundaries, but may have biosocial effects through people’s lived experiences^6^ (**see Box 1**).

Biomarkers that reflect these social disparities are lacking in child populations. Here we test whether DNA-methylation alterations are sensitive to socioeconomic, racial, and ethnic disparities in mental health. Biomarkers developed and validated in adults that associate social disparities with children’s mental health would be a novel tool to (1) improve forecasting of psychiatric illness from childhood to later life, (2) study the early-life social determinants of lifelong mental health, and (3) assess the efficacy of interventions and policies through a new surrogate outcome relevant for later life.

In the current study, we leverage results from *discovery* epigenome-wide studies of adults to create DNA-methylation composite scores that can be used for *prediction* of mental health outcomes and sensitivity to environmental inequality in our independent pediatric sample. Following a preregistered analysis plan (https://osf.io/t3vnp/), two salivary DNA-methylation composite scores and one genetic composite score were selected because their blood-derived composites have been associated with psychiatric symptoms or they have been found to be sensitive to social inequality. First, we examined DNA-methylation profiles of a peripheral proxy for systemic low-grade inflammation (*i.e.,* C-reactive protein, CRP; DNAm-CRP^7^), which in blood samples have previously been found to be associated with internalizing and externalizing symptoms in children^8^. While inflammation is an essential component of immunosurveillance and host defense, a chronic low-grade inflammatory state is a pathological feature of a wide range of chronic health conditions that is also known to affect brain and psychological development^9, 10^. Second, we examined genetic profiles of inflammation (*i.e.,* polygenic scores of CRP, PGS-CRP^11^), because there are genetic correlations of CRP with mood disorders in adults, such as PTSD^12^ and depressive symptoms^13^ that may include causal effects of CRP on depression^14, 15^. Third, we previously reported that socioeconomic disadvantage and Latinx compared to White identity is associated with the pace of multi-system biological aging, as indicated by DunedinPoAm^16^, in an earlier sub-sample of salivary DNA-methylation data from the Texas Twin Project (*N*=600)^17^. In the current analysis, we examine data from 1,183 children and adolescents from the Texas Twin Project aged 8-to-19-years.

### Box 1. Racial and Ethnic Identity, Racism, and Genetic Ancestry.

Racial and ethnic identity, social and institutional racism, and genetic ancestry are not the same thing^18, 19^. Racial and ethnic groups as they are defined in the U.S. were pseudoscientific inventions used as a political tool benefitting White people to justify the enslavement of Africans and genocide of indigenous Americans^4, 5^. Social and institutional racism includes the generational legacy of state-sanctioned social power, resources, representation, and favoritism lived by White people, *i.e.* White privilege, which comes at the cost of marginalized racial and ethnic groups^4, 5^. Racial and ethnic identity is context-specific both in terms of how people self-identify^20^ and how they are categorized by others^21^.

Racial and ethnic disparities in child mental health arise through various factors tied to classism and racism, including inequitable access to high-quality childcare, educational resources, healthcare, nutrition, and differences in exposure to toxicants, family stress, and neighborhood threat, among other factors^22, 23^. Importantly, racial and ethnic identity itself is not a form of adversity or risk factor for mental health, whereas the institutional racial discrimination and chronic nature of interpersonal discrimination associated with racial and ethnic marginalization is^24, 25^.

In contrast, genetic ancestry describes patterns of gene frequencies that people have inherited from their genetic ancestors, and people’s genetic ancestry does not change across social contexts^18, 26^. The lack of biological “reality” underlying race does not mean that race is unimportant: Racial and ethnic identity remains relevant as it pertains to people’s lived experiences, culture, community, social challenges, and opportunity. Thus, these social constructs are not based on “innate” biosocial boundaries, but may have biosocial effects through people’s lived experiences^6^.

Because we are integrating genetic, epigenetic, and social levels of analysis, we employ both labels based on biogeographical ancestry (like “predominantly recent European ancestries”) as relevant to our genomic measure and socially based labels (like “White”) that are more appropriate for population health disparities research^27^.

## Method

### Sample

The Texas Twin Project is an ongoing longitudinal study^28^. Participants in the current study were 1213 children and adolescents that had at least one DNA-methylation sample. 195 participants contributed two DNA-methylation samples (time between repeated samples: *M*=22 months, *SD*=6.5, range 3 to 38 months) and 16 samples were assayed in duplicate for reliability analyses (total methylation sample *n*=1424). After exclusions based on DNA-methylation quality control criteria, data from 1,183 (609 female) children, including 426 monozygotic and 757 dizygotic twins from 611 unique families, aged 8 to 19 years (age M=13.38y, *SD*=2.99y) was analyzed. Participants self-identified as White only (*n*=752), Latinx only (*n*=147), Latinx and White (*n*=97), Black and potentially another race/ethnicity (Black+, *n*=120), Asian and potentially another race/ethnicity (but not Latinx or Black; Asian+, *n*=90), and Indigenous American, Pacific Islander or other (but not Latinx, Black, or Asian; *n*=7). The University of Texas Institutional Review board granted ethical approval.

### Measures

#### DNA-methylation

Saliva samples were collected during a laboratory visit using Oragene kits (DNA Genotek, Canada). The Infinium MethylationEPIC BeadChip kit (Illumina, Inc., USA) was used to assess methylation levels at 850,000 methylation sites. **Supplemental Methods** contain information on preprocessing, **Table 1** a description of DNA-methylation profiles calculation, and **Table 2** descriptive statistics.

**Table 1.**
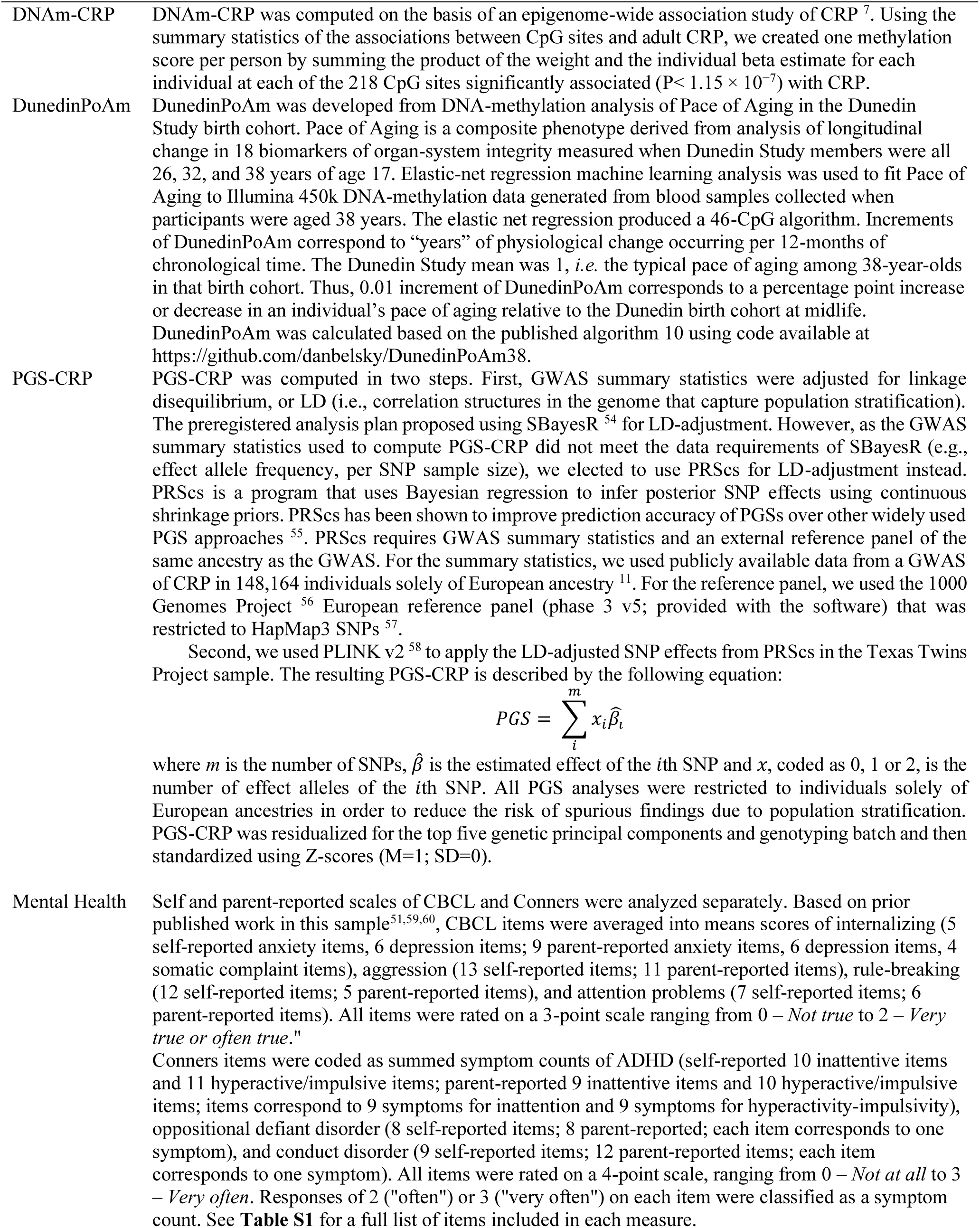
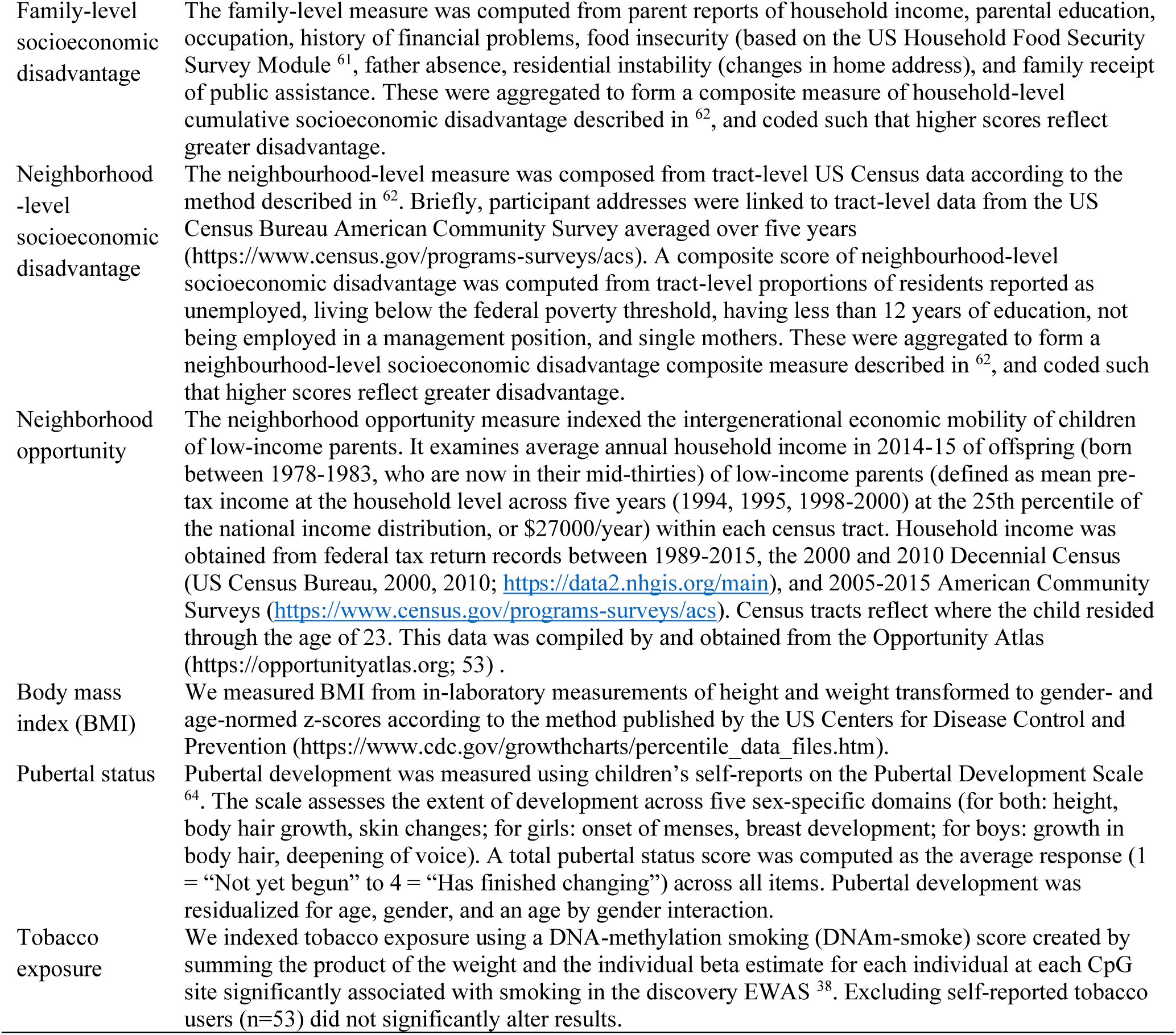
Description of study measures.

**Table 2.** Descriptive statistics.

| Sample | *n* | *M* | *SD* |
| --- | --- | --- | --- |
| DNA<sub>m</sub>-CRP<sup>a</sup> | 1365 | 0.03 | 0.02 |
| DunedinPoAm<sup>a</sup> | 1365 | 1.02 | 0.06 |
| PGS-CRP<sup>b</sup> | 654 | 0 | 1 |
| Parent-reported internalizing | 1311 | 3.05 | 0.39 |
| Parent-reported attention problems | 1311 | 3.27 | 0.50 |
| Parent-reported aggression | 805 | 3.02 | 0.46 |
| Parent-reported rule-breaking | 1311 | 3.14 | 0.70 |
| Parent-reported ADHD | 1266 | 2.39 | 3.87 |
| Parent-reported oppositional defiant disorder | 1266 | 0.63 | 1.39 |
| Parent-reported conduct disorder | 1266 | 0.06 | 0.32 |
| Child-reported internalizing | 1255 | 0.55 | 0.40 |
| Child -reported attention problems | 1255 | 0.72 | 0.43 |
| Child -reported aggression | 1255 | 0.39 | 0.27 |
| Child -reported rule-breaking | 1255 | 0.25 | 0.24 |
| Child -reported ADHD | 1104 | 4.45 | 3.99 |
| Child -reported oppositional defiant disorder | 1104 | 1.06 | 1.27 |
| Child -reported conduct disorder | 1104 | 0.11 | 0.46 |
| Family-level socioeconomic disadvantage | 993 | -0.02 | 0.96 |
| Neighborhood-level socioeconomic disadvantage | 1218 | -0.02 | 1 |
| Neighborhood opportunity | 950 | 0.31 | 0.62 |
| Body mass index | 1364 | 0.4 | 1.34 |
| Pubertal development | 1325 | 2.61 | 0.92 |
| Tobacco use (yes/no) | 58/631 | – | – |
| DNA<sub>m</sub>-smoke | 1365 | -13.62 | 1.84 |

<sup>a</sup> After exclusion of participants based on DNA-methylation preprocessing (n=44), excluding technical replicates (n=15), and including repeated samples (n= 182). Means of raw scores before residualizing for cell composition, array, slide, and batch. Scores were standardized (mean = 0, SD = 1) for analyses.
<sup>b</sup> PGS-CRP only computed for individuals solely of recent European ancestries.

#### Genetics

Genetic data was used to calculate a polygenic score for CRP (PGS-CRP; description in **Table 1**). The PGS-CRP is an approximate indicator of an individual’s genetic liability for developing high levels of CRP^26^. All PGS-CRP analyses were restricted to individuals solely of European ancestries in order to reduce the risk of spurious findings due to population stratification^26^. **Supplemental Methods** contain information on genotyping.

#### Mental health

Multiple dimensions of child and adolescent mental health were measured using self-reports and parent-reports on abbreviated versions of the Achenbach Child Behavior Checklist^29, 30^, and the *DSM-IV* symptom count scales of the Conners 3^31^. **Table 1** contains a description of mental health composite measure calculation and **Table S1** a full list of items included in each measure.

#### Socioeconomic context

We measured children’s socioeconomic disadvantage at the family and neighborhood levels of analysis (**Table 1**).

#### Developmental covariates and tobacco exposure

Body mass index^32^ (BMI), pubertal status^33, 34^, and tobacco exposure^35^ have been associated with early life disadvantage as well as differential DNA-methylation patterns^36–38^. We therefore consider these factors in our analysis as covariates (**Table 1**).

### Statistical analyses

Fourteen mental health outcomes were examined in each model, including 7 self-reported and 7 parent-reported aggregate measures of (1) internalizing, (2) attention problems, (3) aggression, (4) rule-breaking, (5) ADHD, (6) oppositional defiant disorder (ODD), and (7) conduct disorder (CD).

We performed multilevel, multivariate regression models fit with FIML in *Mplus* 8.2 software^39^. All models included a random intercept, representing the family-level intercept of the dependent variable, to correct for non-independence of twins. To account for nesting of repeated measures within individuals, a sandwich correction was applied to the standard errors in all analyses. All models included age, gender, and an age by gender interaction as covariates. We used the Benjamini-Hochberg false discovery rate method (FDR)^40^ to account for multiple testing. We report standardized effect size estimates and nominal *P*-values as significant, when the FDR-corrected *P*-values were below alpha<0.05.

## Results

### Mental health is socially stratified in children

We observed socioeconomic disparities in parent- and child-reported psychiatric burden. After correcting for seven comparisons, family-level disadvantage was associated with more parent-reported internalizing, attention problems, aggression, rule-breaking, ADHD, and ODD symptoms, but not CD symptoms (effect sizes *r*=0.11 to 0.26; **Figure 1** and **Supplemental Table S2)**. Family-level disadvantage was also associated with higher children’s self-reported attention problems, aggression, and rule-breaking (effect sizes *r*=0.16 to 0.24; **Figure 1** and **Supplemental Table S3)**.

**Figure 1.**
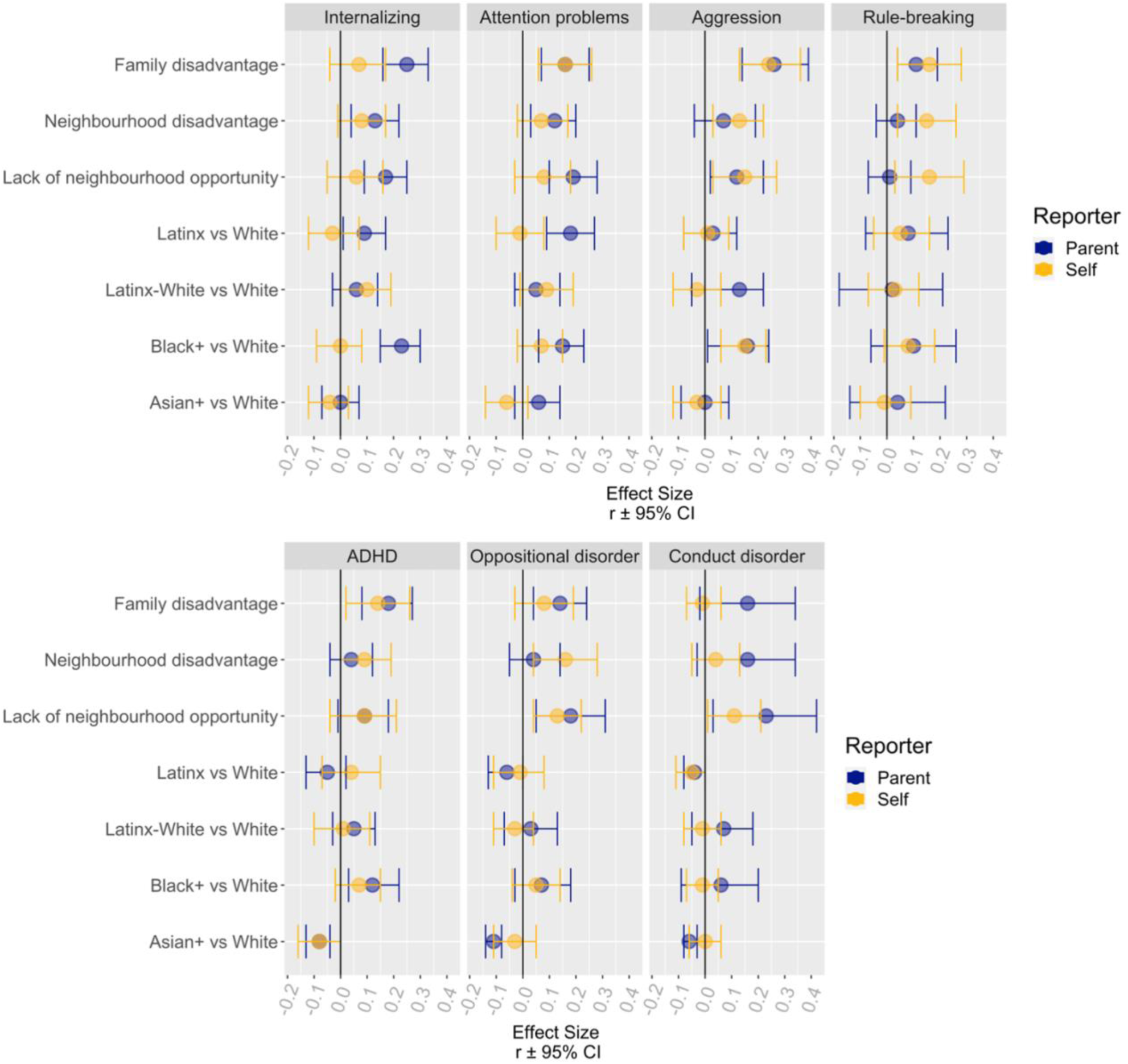
Associations between measures of socioeconomic inequality, racial and ethnic identity with parent- and self-reported mental health in children and adolescents. Depicted are standardized effect size estimates and 95% confidence intervals. See Supplemental Table S2 and S3 for coefficients.

Neighbourhood-level disadvantage was associated with higher parent-reported internalizing (*r*=0.13, CI=0.04 to 0.22, *P*=0.003) and attention problems (*r*=0.12, CI=0.03 to 0.20, *P*=0.008). Neighborhood opportunity was associated with lower parent-reported internalizing, attention problems, aggression, and ODD (effect sizes *r*=-0.12 to -0.19; **Figure 1** and **Table S2)**. Neighbourhood-level disadvantage and less neighbourhood opportunity were both also associated with higher child-reported aggression, rule-breaking, and ODD (effect sizes *r*=0.13 to 0.16 and *r*=-0.13 to -0.16, respectively; **Figure 1** and **Table S3)**.

We also observed racial/ethnic disparities in parent- and child-reported mental health. Children identifying as Latinx-only or Black+ had higher parent-reported internalizing and attention problems than White-only identifying children (effect sizes *b*=0.09 to 0.23; **Figure 1** and **Table S2)**. Black+ identifying children also showed more parent-reported symptoms of ADHD. Asian+ identifying children had lower parent-reported ADHD, ODD, and CD than White-only children (effect sizes *b*=-0.06 to -0.11). Child-reported mental health was largely unrelated to their race/ethnicity except that Black+ identifying children self-reported higher rates of aggression than White-only children (*b*=0.15, CI=0.06 to 0.23, *P=*0.001).

We then examined the degree to which socioeconomic inequality statistically accounted for racial/ethnic disparities in mental health. Children reporting Black+ and Latinx-only identities lived in substantially more socioeconomically disadvantaged families and neighborhoods compared to children reporting White-only identity (effect sizes *b*=0.25 to 0.43; **Table S4**). Family-level disadvantage accounted for differences between Latinx-only and White-only children’s parent-reported internalizing, but not attention problems (**Table S2)**. Family-level disadvantage accounted for differences between Black+ and White-only children’s parent-reported ADHD and child-reported aggression, but not differences in parent-reported internalizing or attention problems (**Table S2 and S3)**. After accounting for family-level disadvantage, Latinx-only children self-reported lower rates of CD than White-only children.

Linear regression analysis (not preregistered) indicated that White identity compared to racial/ethnic categories associated with marginalization was associated with less parent-reported internalizing (*r=*-0.16, 95% CI=-0.23 to -0.08, *P*<0.001) and fewer attention problems (*r=*-0.19, CI=-0.27 to -0.11, P<0.001), but not less aggression (*r=*-0.09, CI=-0.19 to 0.01, *P=*0.079), rule-breaking (*r=*-0.05, CI=-0.13 to 0.02, *P*=0.139), ADHD (*r=*-0.01, CI=-0.09 to 0.07, *P*=0.796), ODD (*r=*0.03, CI=-0.05 to 0.11, *P*=0.471), or CD (*r=*0.00, CI=-0.13 to 0.13, *P*=0.990). Though attenuated, White identity remained a statistically significant predictor of parent-reported internalizing (*r=*-0.09, CI=-0.16 to -0.02, *P*=0.012) and attention problems (*r=*-0.15, CI=-0.38 to -0.19, *P*<0.001) after accounting for family-level disadvantage (racial/ethnic identity and family-level disadvantage: *r=*-0.29, CI=-0.38 to -0.19, *P*<0.001). White identity also remained a significant predictor of parent-reported internalizing (*r=*-0.09, CI=-0.16 to -0.01, *P*=0.020) and attention problems (*r=*-0.15, CI=-0.23 to -0.06, *P*=0.001), after accounting for both family- and neighborhood-level socioeconomic disadvantage together (*r=*-0.34, CI=-0.42 to -0.26, *P*<0.001).

### Salivary DNA-methylation profiles are socially stratified in children

Analyses of 15 technical replicates (one sample did not pass quality control) suggested moderate-to-good reliability of DNA-methylation profiles residualized for technical artifacts and cell composition (ICC for DNAm-CRP=0.73, DunedinPoAm=0.84; see **Table 1** for descriptive statistics). Biometric models using the twin family structure, where the sum of additive genetic factors (*A*) and environmental factors shared by twins living in the same home (*C*) represents a lower bound estimate of reliability (because it does not include measurement error), also suggested good reliability of DNA-methylation profiles (*A*+*C* variation for DNAm-CRP= 60.7%, DunedinPoAm= 54.2%, accounting for age and gender).

Higher DNAm-CRP was correlated with higher DunedinPoAm (*r* =0.89, CI=0.81 to 0.96, *P*<0.001, accounting for age and gender). This is not surprising, as CRP levels were one of the 18 biomarkers that the pace of aging algorithm was trained on, and 7 CpG sites overlapped across measures^16^.

Older children had higher DNAm-CRP (*r* =0.35, CI=0.26 to 0.44, *P*<0.001) and DunedinPoAm profiles (*r* =0.13, CI=0.02 to 0.23, *P*=0.018). Boys had lower DNAm-CRP (*b*=- 0.26, CI=-0.34 to -0.18, *P*<0.001) and DunedinPoAm (*b*=-0.18, CI=-0.27 to -0.10, *P*<0.001) profiles (*b*=0.06, CI= -0.02 to 0.14, *P*=0.143) as compared to girls. All models included age, gender, and an age by gender interaction as covariates.

We observed salivary DNA-methylation profiles that differed by socioeconomic and racial/ethnic inequality. As reported in Raffington et al.^41^, children from socioeconomically disadvantaged families, socioeconomically disadvantaged neighborhoods, neighborhoods with less neighborhood opportunity, and Latinx-only or Black+ identity relative to White-only identity exhibited DNA-methylation profiles associated with higher chronic inflammation and a faster pace of biological aging (effect sizes *r*=0.08 to 0.28; **Table S5**). The analyses of Raffington et al.^41^ were preregistered at the same time as the preregistration of the present study. See **Table S6** for effect size estimates between socioeconomic inequality and DNA-methylation profiles reported separately for each racial/ethnic group (not preregistered).

### Salivary DNA-methylation profiles are associated with child and adolescent mental health

We assessed whether salivary DNA-methylation profiles were associated with child mental health. Higher DNAm-CRP was associated with higher parent-reported internalizing (*r*=0.15, CI=0.05 to 0.25, *P*=0.004) and aggression (*r*=0.17, CI=0.04 to 0.31, *P*=0.013; **Figure 2** and **Table S7**). Faster DunedinPoAm was also associated with higher parent-reported internalizing (*r*=0.15, CI=0.05 to 0.25, *P*=0.002).

**Figure 2.**
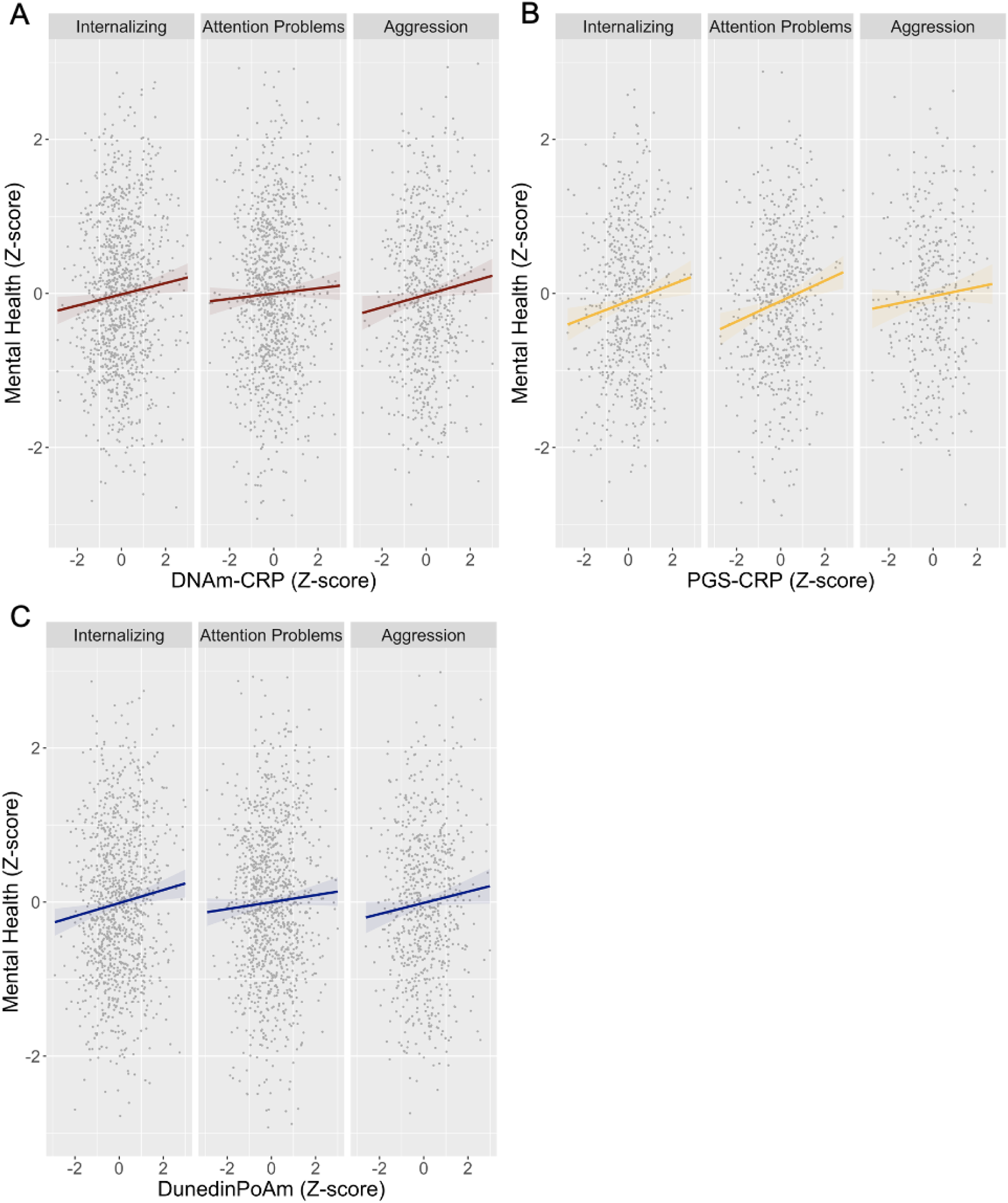
Associations between A) DNAm-CRP, B) PGS-CRP, and C) DunedinPoAm with parent-reported mental health in children and adolescents. DNA-methylation profiles and mental health values are in standard deviation units. Higher DNAm-CRP values indicate a methylation profile associated with higher chronic inflammation. Higher PGS-CRP values indicate a genetic profile associated with higher chronic inflammation. Higher DunedinPoAm values indicate a methylation profile associated with faster biological aging. See **Supplemental Table S7** for standardized regression coefficients.

Since both profiles were associated with parent-reported internalizing, we performed commonality analyses to examine the proportion of overlapping and unique variation explained. DNAm-CRP and DunedinPoAm explained largely unique variation in parent-reported internalizing (DNAm-CRP alone: 2.5%, DunedinPoAm alone: 2.2%, combined: 4.5%).

DNA-methylation profiles were not associated with children’s self-reported mental health (**Table S8**). See **Table S9 and S10** for effect size estimates between DNA-methylation profiles and mental health reported separately for each racial/ethnic group (not preregistered).

### DNA-methylation partially accounts for associations of family-level disadvantage and racial and ethnic marginalization with mental health

We observed that DNAm-CRP and DunedinPoAm partially accounted for associations of family-level disadvantage and racial/ethnic disparities in mental health. DNAm-CRP, but not DunedinPoAm, partially mediated family-level disadvantage on internalizing (indirect effect 16% of total effect), but not on aggression. Both DNAm-CRP and DunedinPoAm partially mediated the impact of being racially minoritized compared to not on internalizing (indirect DNAm-CRP effect 12% of total effect; indirect DunedinPoAm effect 19% of total effect).

### BMI statistically accounts for associations of DNA-methylation with mental health

We next examined the role of BMI, pubertal status, and DNAm-smoke in associations of DNA-methylation with mental health (see **Table S7**). Associations of DNAm-CRP and DunedinPoAm with mental health were largely accounted for by controlling for BMI, but not by controlling for pubertal status. Associations of DNAm-CRP with aggression were largely accounted for by controlling for DNAm-smoke.

### Within-family analyses indicate family-level stratification of DNA-methylation

We further assessed the extent to which DNA-methylation associations with mental health are robust to complete genetic and family-level environmental control in a bivariate ACE model that used the twin family structure. We found no evidence to suggest that monozygotic twins who differ from their co-twins in DNA-methylation show corresponding differences in their mental health (see **Table S11**). This is consistent with the hypothesis that DNA-methylation associations with mental health represents (potentially partially unmeasured) effects of family-level stratification.

### Polygenic scores of CRP are associated with mental health

In an analysis restricted to participants solely of European genetic ancestry, PGS-CRP was not correlated with DNAm-CRP (*r*=0.04, CI=-0.08 to 0.17, *P*=0.476) or DunedinPoAm (*r*=- 0.04, CI=-0.18 to 0.10, *P*=0.563).

Higher PGS-CRP was associated with higher parent-reported internalizing (*r*=0.14, CI 0.04 to 0.23, *P*=0.004) and attention problems (*r*=0.16, CI=0.05 to 0.28, *P*=0.004; **Figure 1** and **Table S7**) and these associations were unaffected by controlling for BMI, pubertal status, DNAm-smoke, and family-level disadvantage. PGS-CRP was not associated with children’s self-reported mental health (**Table S8**).

Commonality analyses showed that PGS-CRP explained largely unique variation in parent-reported internalizing (PGS-CRP alone: 2%) relative to both DNAm-CRP (combined: 4.5%) and DunedinPoAm (combined: 4.2%).

In exploratory preregistered analyses of 364 dizygotic twin pairs, PGS-CRP did not account for differences in internalizing (*r=*0.07, 95% CI=-0.06 to 0.20, *P*=0.316) or attention problems (*r=*0.13, 95% CI=-0.02 to 0.27, *P*=0.094), although we caution that effect size estimates warrant a repeated examination of this question in a larger sample.

## Discussion

We analyzed epigenetic data from 1183 children and adolescents participating in the Texas Twin Project to examine whether salivary DNA-methylation patterns are socially stratified and are associated with mental health. We find that salivary DNA-methylation profiles of inflammation (DNAm-CRP), faster pace of biological aging (DunedinPoAm) and parent-reported child and adolescent mental health, consistently differed by socioeconomic inequality and self-reported racial/ethnic identity. Additionally, these salivary DNA-methylation profiles and genetic profiles of inflammation (PGS-CRP) were associated with children’s internalizing, aggression, and attention problems. Moreover, salivary DNA-methylation profiles partially accounted for child mental health disparities related to family-level disadvantage, racial, and ethnic identities.

Our findings linking environmental inequality to epigenetic profiles and mental health were most consistent for internalizing symptoms, which is in line with previous research on peripheral inflammation^42^ and a blood-based study on DNA-methylation of inflammation in children^8^. These results add to a growing body of research in animals and humans that links exposure to social adversity, DNA-methylation changes, and gene expression markers associated with inflammation^10, 43, 44^. Animal research suggests that inflammation affects synaptogenesis, synaptic survival, and myelination, for instance in neural circuitries subserving executive control, and threat and reward processing that are pertinent to emotional and behavioral regulation^45^. Further, inflammation and DNA-methylation alterations are environmentally-sensitive hallmarks of biological aging^46–48^. We found that body mass index largely accounted for associations of DNAm-CRP and DunedinPoAm with mental health, which corresponds to new insights casting obesity as an inflammatory disease of accelerated biological aging^49^.

Highlighting the distinctly harmful role of social and institutional racism, we found that even after accounting for the lower rates of socioeconomic disadvantage experienced by White children, White identifying children sustained a lower burden of mental health symptoms. Thus, socioeconomic inequality captures some but not all of the pathways through which social and institutional racism impacts children’s mental health^24^. Moreover, our categorization of children’s complex racial and ethnic identities that for instance grouped Black and Black-White identifying children together is likely to dilute and underestimate the bio-psychological effects of racial marginalization as the proximity to White privilege and experiences of marginalization are not the same between these identities.

Further research is needed to contextualize the biological and mental health effects of race-based discrimination, prejudice, and oppression experienced by marginalized youth compared to the favoritism and privilege experienced by Whites^3, 6, 50^. While our findings are in line with the hypothesis that DNA-methylation alterations are one mechanism for how psychosocial stress is biologically embedded to injure children’s psychiatric health, our observational design is, of course, not well-suited to evaluate mechanistic hypotheses. Randomized controlled trials and natural experiments that involve exogenous changes to various aspects of social inequality (*e.g.,* eliminating redlining, child tax credits) are necessary to establish whether children’s salivary DNA-methylation profiles change in response to intervention in a way that is tied to mental health.

In contrast to results seen for parent-reported psychiatric symptoms, child-reported mental health was generally not associated with DNA-methylation or PGS-CRP profiles. Previous research in this sample has found that children and parents agree minimally on specific symptoms scales^51^. In addition, both children and parent self-reports can be influenced by their level of insight of their specific psychopathological burden, and the degree to which they have internalised narratives associated with being racially minoritized, *i.e,* internalised racism^52^. We further acknowledge that restricting PGS-CRP analyses to participants solely of European genetic ancestry is a major study limitation. Future research should explore whether recent approaches to compute ancestry deconvoluted polygenic scores^53^ can be applied to PGS-CRP and validated with serum CRP in ancestrally diverse samples.

In conclusion, our findings suggest that salivary DNA-methylation patterns of higher inflammation and faster pace of biological aging are economically and racially stratified and are associated with child mental health. Because saliva can easily be collected in large-scale pediatric epidemiological studies and may indicate emerging health conditions, these DNA-methylation profiles can be employed in research seeking to understand and prevent economic and racial disparities in childhood mental health.

## Data Availability

Participants are drawn from a unique population (twins) from a tightly-defined geographic region. In addition, participants are sampled from a vulnerable population (children), many of whom are ethnic minorities and/or live in low income contexts. Finally participants provide highly sensitive psychological, economic, academic, and genetic information. Thus because of the vulnerable status of many participants in the sample, the strong potential for deductive identification, and the sensitive nature of the information collected, data from the Texas Twin Project are not shared with individuals outside of the research team.

## Acknowledgment Section

### Funding

We gratefully acknowledge all participants of the Texas Twin Project. This research was supported by National Institutes of Health (NIH) grants R01HD083613 and R01HD092548. LR was supported by the German Research Foundation (DFG). MWP was supported by a training grant from NIMH, T32MH015442. ZA is supported by a Marie Skłodowska-Curie Fellowship from the European Union (894675). KPH and EMTD are Faculty Research Associates of the Population Research Center at the University of Texas at Austin, which is supported by a NIH grant P2CHD042849. EMTD is a member of the Center on Aging and Population Sciences (CAPS) at The University of Texas at Austin, which is supported by NIH grant P30AG066614. KPH and EMTD were also supported by Jacobs Foundation Research Fellowships. LR and KPH had full access to all the data in the study and take responsibility for the integrity of the data and the accuracy of the data analysis.

### Conflicts of interest

Not applicable.

### Ethics approval

The University of Texas at Austin Institutional Review board granted ethical approval.

### Consent to participate

Informed consent to participate in the study was obtained from all participants and their parent or legal guardian.

### Consent for publication

Not applicable.

### Availability of data and material

Because of the high potential for deductive identification in this special population from a geographically circumscribed area, and the sensitive nature of information collected, data from the Texas Twin Project are not shared with individuals outside of the research team.

### Code availability

Code will be shared by the first author upon request.

### Author Contributions

KPH, EMTD, & LR developed the study concept and design. LR, PT, LV, & TM performed the data analysis under the supervision of EMTD and KPH. LR drafted the manuscript. All authors provided critical revisions to the analysis plan and manuscript and approved the final version of the manuscript for submission.

## Supplemental Methods

### DNA-methylation preprocessing

DNA extraction and methylation profiling was conducted by Edinburgh Clinical Research Facility (UK). DNA-methylation preprocessing was primarily conducted with the ‘minfi’ package in R ^65^. Within-array normalization was performed to address array background correction, red/green dye bias, and probe type I/II correction, and it has been noted that at least part of the probe type bias is a combination of the first two factors ( . Noob preprocessing as implemented by minfi’s “preprocessNoob” ^66^ is a background correction and dye-bias equalization method that has similar within-array normalization effects on the data as probe type correction methods such as BMIQ (Teschendorff et al., 2013).

In line with our preregistered preprocessing plan, CpG probes with detection p > 0.01 and fewer than 3 beads in more than 1% of the samples and probes in cross-reactive regions were excluded ^67^. None of these failed probes overlapped with the probes used for DNA-methylation scores. 44 samples were excluded because (1) they showed low intensity probes as indicated by the log of average methylation <9 and their detection *p*-value was > 0.01 in >10% of their probes, (2) their self-reported and methylation-estimated sex mismatch, and/or (3) their self-reported and DNA-estimated sex mismatch. Cell composition of immune and epithelial cell types (*i.e.,* CD4+ T-cell, natural killer cells, neutrophilseosinophils, B cells, monocytes, CD8+ T-cell, and granulocytes) were estimated using a newly developed child saliva reference panel implemented in the R package “BeadSorted.Saliva.EPIC” within “ewastools” ^68^. Surrogate variable analysis was used to correct methylation values for batch effects using the “combat” function in the SVA package ^69^.

### Genotyping and imputation

DNA samples were genotyped at the University of Edinburgh using the Illumina Infinium PsychArray, which assays ∼590,000 single nucleotide polymorphisms (SNPs), insertions-deletions (indels), copy number variants (CNVs), structural variants, and germline variants across the genome. Genetic data was subjected to quality control procedures recommended for chip-based genomic data ^70, 71^. Briefly, samples were excluded on the basis of poor call rate (< 98%) or inconsistent self-reported and biological sex, while variants were excluded if missingness exceeded 2%. As further variant-level filtering has been shown to have a detrimental effect on imputation quality ^72^, quality control thresholds for minor allele frequency (MAF) and Hardy–Weinberg equilibrium (HWE) were applied after phasing and imputation.

Untyped markers were imputed on the Michigan Imputation Server (https://imputationserver.sph.umich.edu). Specifically, genotypes were phased and imputed with Eagle v2.4 and Minimac4 (v1.5.7), respectively, while using the 1000 Genomes Phase 3 v5 reference panel ^56^. To ensure that only high-quality typed and imputed markers were used for analysis, variants were excluded if they had a MAF < 1e-3, a HWE p-value < 1e-6, or an imputation quality score < .90. These procedures produced a final set of 4,703,309 genetic markers to be used in analyses.

## Supplemental Tables

**Table S1.**
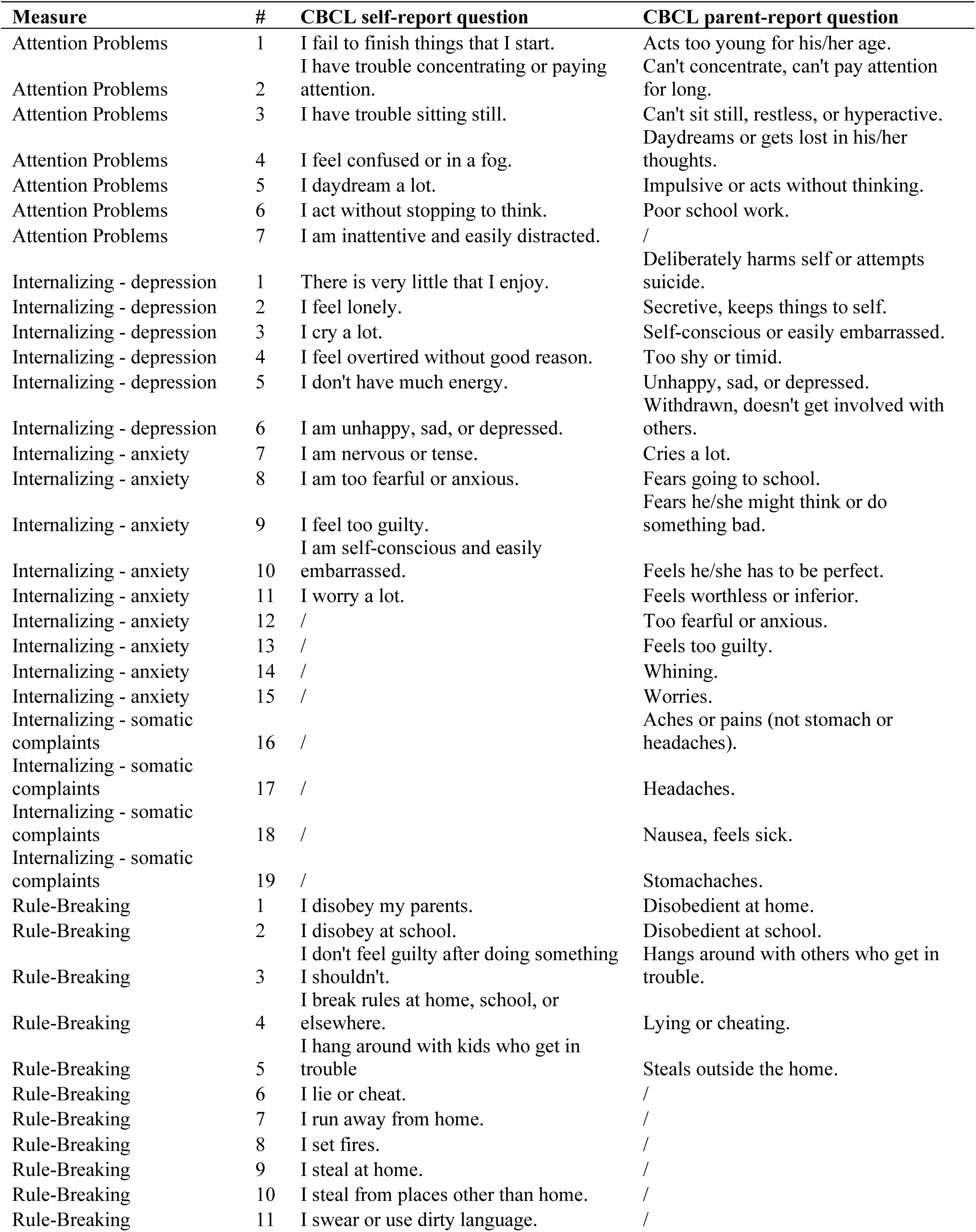
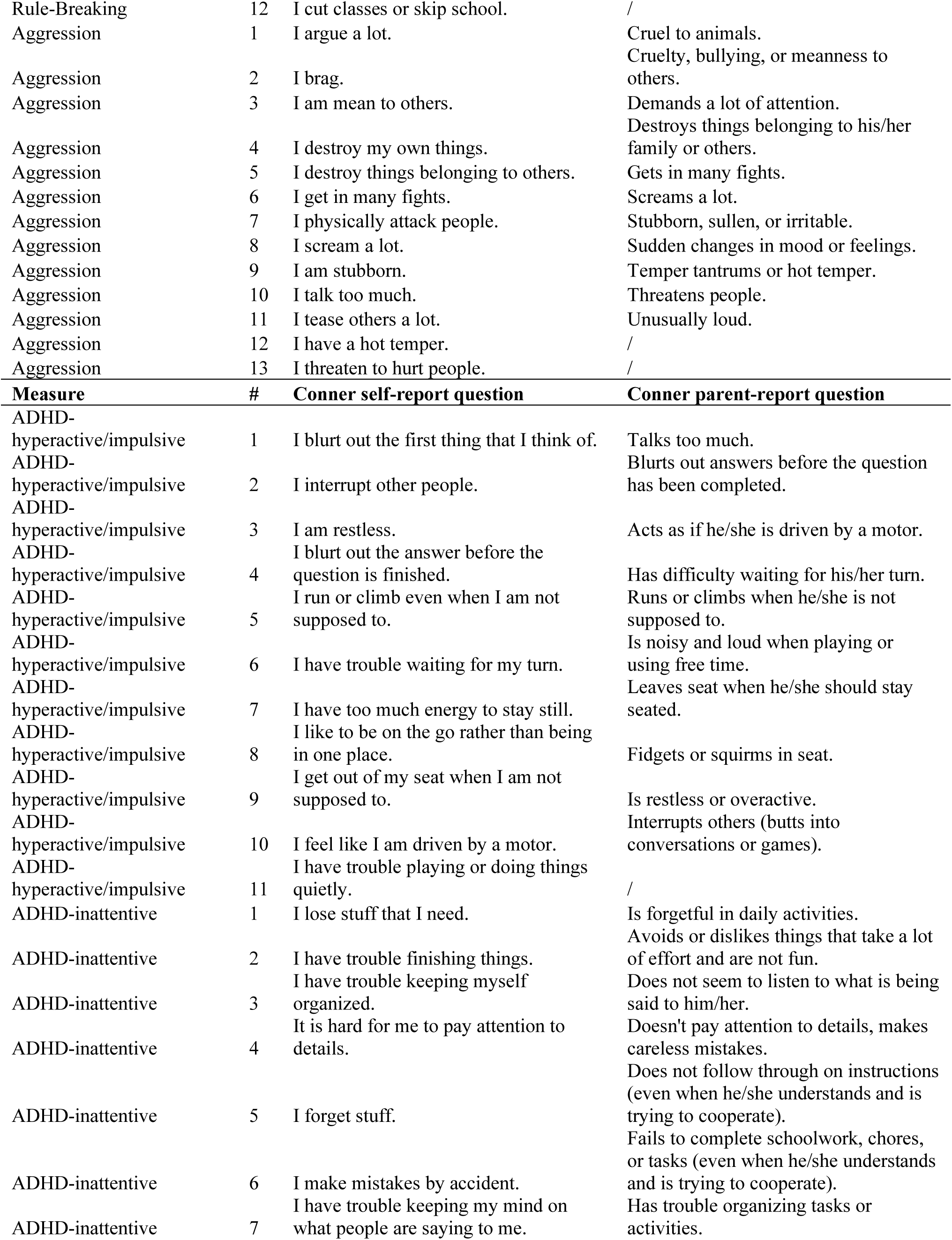
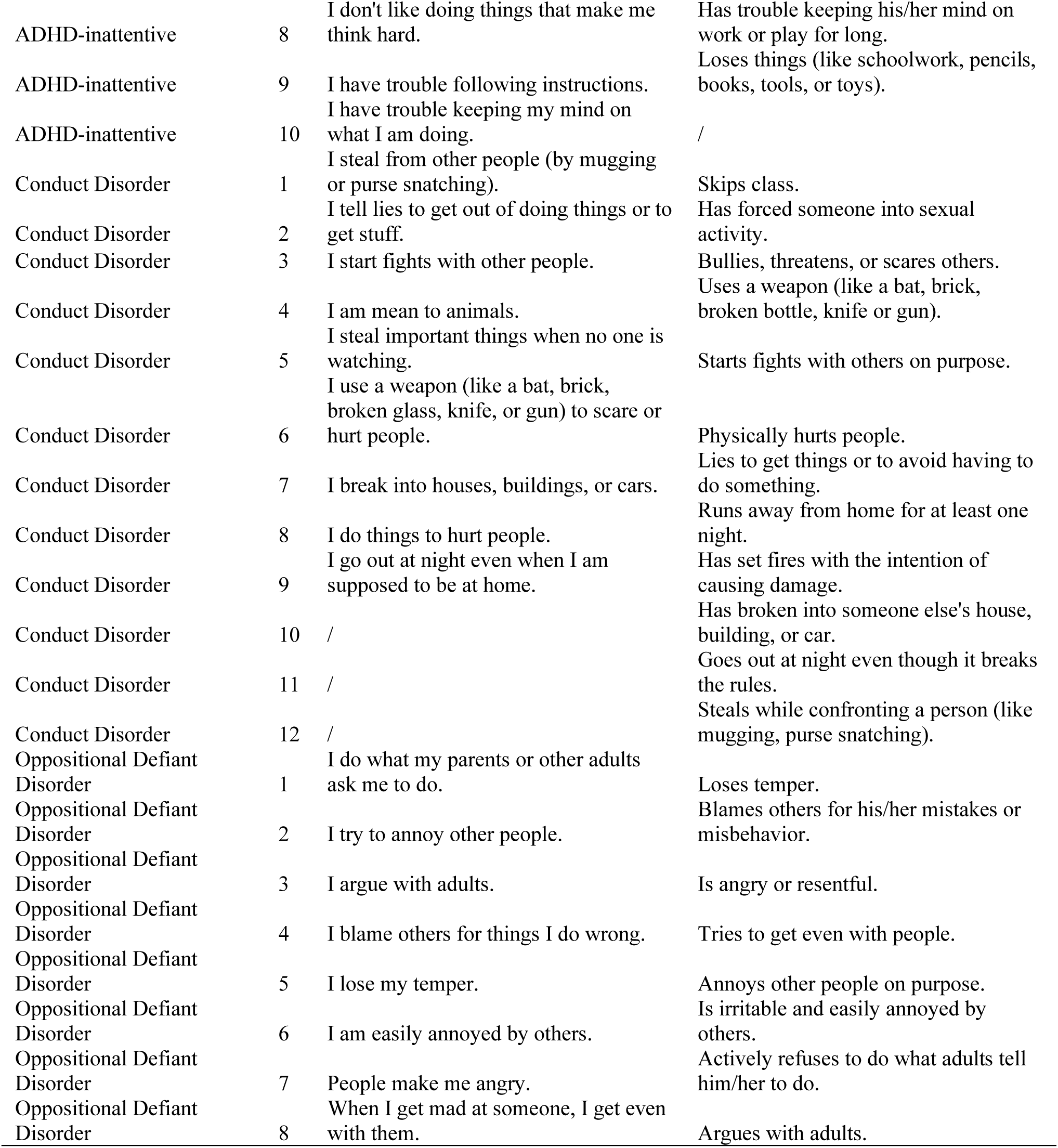
CBCL and Conners items included in each mental health summary measure.

**Table S2.** Associations between socioeconomic inequality and racial/ethnic identity with parent-reported mental health.

|  | Internalizing |  |  | Attention Problems |  |  | Aggression |  |  | Rule-Breaking |  |  | ADHD |  |  | ODD |  |  | CD |  |  |
| --- | --- | --- | --- | --- | --- | --- | --- | --- | --- | --- | --- | --- | --- | --- | --- | --- | --- | --- | --- | --- | --- |
|  | No further covariates<sup>a</sup> |  |  |  |  |  |  |  |  |  |  |  |  |  |  |  |  |  |  |  |  |
|  | *b* | CI | *p* | *b* | CI | *p* | *b* | CI | *p* | *b* | CI | *p* | *b* | CI | *p* | *b* | CI | *p* | *b* | CI | *p* |
| Family-level disadvantage | 0.25 | 0.16, 0.33 | **0.000** | 0.16 | 0.07, 0.25 | **0.001** | 0.26 | 0.14, 0.39 | **0.000** | 0.11 | 0.04, 0.19 | **0.004** | 0.18 | 0.08, 0.27 | **0.000** | 0.14 | 0.04, 0.24 | **0.005** | 0.16 | -0.02, 0.34 | 0.088 |
| Neighborhood-level disadvantage | 0.13 | 0.04, 0.22 | **0.003** | 0.12 | 0.03, 0.20 | **0.008** | 0.07 | -0.04, 0.19 | 0.219 | 0.04 | -0.04, 0.11 | 0.364 | 0.04 | -0.04, 0.12 | 0.286 | 0.04 | -0.05, 0.14 | 0.365 | 0.16 | -0.03, 0.34 | 0.097 |
| Neighborhood opportunity | -0.17 | -0.25, -0.09 | **0.000** | -0.19 | -0.28, -0.10 | **0.000** | -0.12 | -0.22, -0.02 | 0.016 | -0.01 | -0.09, 0.07 | 0.826 | -0.09 | -0.18, 0.01 | 0.068 | -0.18 | -0.31, -0.05 | **0.008** | -0.23 | -0.42, -0.03 | 0.022 |
| Latinx | 0.09 | 0.01, 0.17 | **0.021** | 0.18 | 0.09, 0.27 | **0.000** | 0.03 | -0.08, 0.12 | 0.689 | 0.08 | -0.08, 0.23 | 0.352 | -0.05 | -0.13, 0.02 | 0.168 | -0.06 | -0.13, 0.00 | 0.060 | -0.04 | -0.08, 0.00 | 0.047 |
| Latinx-White | 0.06 | -0.03, 0.14 | 0.218 | 0.05 | -0.03, 0.14 | 0.213 | 0.13 | -0.05, 0.22 | 0.232 | 0.02 | -0.18, 0.21 | 0.854 | 0.05 | -0.03, 0.13 | 0.250 | 0.03 | -0.07, 0.13 | 0.570 | 0.07 | -0.05, 0.18 | 0.254 |
| Black+ | 0.23 | 0.15, 0.30 | **0.000** | 0.15 | 0.06, 0.23 | **0.001** | 0.16 | 0.01, 0.24 | 0.027 | 0.10 | -0.06, 0.26 | 0.204 | 0.12 | 0.03, 0.22 | **0.009** | 0.07 | -0.03, 0.18 | 0.173 | 0.06 | -0.09, 0.20 | 0.399 |
| Asian+ | 0.00 | -0.07, 0.07 | 0.946 | 0.06 | -0.03, 0.14 | 0.173 | 0.00 | -0.09, 0.09 | 0.995 | 0.04 | -0.14, 0.22 | 0.647 | -0.08 | -0.13, -0.04 | **0.001** | -0.11 | -0.14, -0.08 | **0.000** | -0.06 | -0.08, -0.03 | **0.001** |
|  | Controlling for family-level disadvantage |  |  |  |  |  |  |  |  |  |  |  |  |  |  |  |  |  |  |  |  |
|  | *b* | CI | *p* | *b* | CI | *p* | *b* | CI | *p* | *b* | CI | *p* | *b* | CI | *p* | *b* | CI | *p* | *b* | CI | *p* |
| Family-level disadvantage | 0.17 | 0.07, 0.26 | **0.000** | 0.09 | -0.02, 0.19 | 0.105 | 0.24 | 0.11, 0.37 | **0.000** | 0.13 | 0.03, 0.22 | **0.008** | 0.16 | 0.05, 0.27 | **0.004** | 0.16 | 0.05, 0.26 | **0.004** | 0.14 | -0.01, 0.29 | 0.063 |
| Latinx | 0.07 | -0.02, 0.16 | 0.120 | 0.16 | 0.06, 0.26 | **0.002** | -0.02 | -0.15, 0.11 | 0.774 | -0.01 | -0.11, 0.10 | 0.909 | -0.09 | -0.18, 0.01 | 0.078 | -0.11 | -0.20, -0.03 | **0.007** | -0.10 | -0.17, -0.03 | **0.005** |
| Latinx-White | 0.06 | -0.04, 0.15 | 0.239 | 0.05 | -0.04, 0.15 | 0.283 | 0.13 | -0.03, 0.28 | 0.109 | -0.02 | -0.10, 0.07 | 0.692 | 0.02 | -0.08, 0.12 | 0.685 | 0.03 | -0.09, 0.14 | 0.682 | 0.07 | -0.08, 0.22 | 0.360 |
| Black+ | 0.16 | 0.06, 0.25 | **0.002** | 0.14 | 0.03, 0.26 | **0.017** | 0.09 | -0.06, 0.23 | 0.250 | -0.01 | -0.12, 0.10 | 0.844 | 0.06 | -0.05, 0.16 | 0.286 | -0.02 | -0.13, 0.09 | 0.732 | 0.09 | -0.12, 0.30 | 0.410 |
| Asian+ | -0.03 | -0.11, 0.04 | 0.433 | 0.08 | -0.02, 0.18 | 0.133 | 0.02 | -0.1, 0.13 | 0.773 | 0.04 | -0.06, 0.14 | 0.421 | -0.07 | -0.13, -0.01 | **0.016** | -0.10 | -0.14, -0.06 | **0.000** | -0.05 | -0.11, 0.00 | 0.075 |

Standardized regression coefficients (*b*) and 95% confidence intervals (CIs) and uncorrected *p*-value calculated by regressing parent-reported mental health on socioeconomic inequality and racial/ethnic identity (with and without controlling for family-level disadvantage). *P*-values, where FDR corrected *p*-values < 0.05, are marked in bold. White-only identity is reference group. <sup>a</sup>All models included covariate adjustment for age, gender, and an age by gender interaction. *Note:* ADHD=attention-deficit hyperactivity disorder; ODD=oppositional defiant disorder; CD= conduct disorder.

**Table S3.** Associations between socioeconomic inequality and racial/ethnic identity with self-reported mental health.

|  | Internalizing |  |  | Attention Problems |  |  | Aggression |  |  | Rule-Breaking |  |  | ADHD |  |  | ODD |  |  | CD |  |  |
| --- | --- | --- | --- | --- | --- | --- | --- | --- | --- | --- | --- | --- | --- | --- | --- | --- | --- | --- | --- | --- | --- |
|  | *b* | CI | *p* | *b* | CI | *p* | *b* | CI | *p* | *b* | CI | *p* | *b* | CI | *p* | *b* | CI | *p* | *b* | CI | *p* |
|  | No further covariates<sup>a</sup> |  |  |  |  |  |  |  |  |  |  |  |  |  |  |  |  |  |  |  |  |
| Family-level disadvantage | 0.07 | -0.04, 0.17 | 0.237 | 0.16 | 0.06, 0.26 | **0.001** | 0.24 | 0.13, 0.36 | **0.000** | 0.16 | 0.04, 0.28 | **0.010** | 0.14 | 0.02, 0.26 | 0.024 | 0.08 | -0.03, 0.19 | 0.142 | -0.01 | -0.07, 0.06 | 0.778 |
| Neighborhood-level disadvantage | 0.08 | -0.01, 0.17 | 0.085 | 0.07 | -0.02, 0.17 | 0.117 | 0.13 | 0.03, 0.22 | **0.008** | 0.15 | 0.04, 0.26 | **0.009** | 0.09 | 0.00, 0.19 | 0.046 | 0.16 | 0.04, 0.28 | **0.009** | 0.04 | -0.05, 0.13 | 0.409 |
| Neighborhood opportunity | -0.06 | -0.16, 0.05 | 0.280 | -0.08 | -0.18, 0.03 | 0.171 | -0.15 | -0.27, -0.03 | **0.013** | -0.16 | -0.29, -0.03 | **0.017** | -0.09 | -0.21, 0.04 | 0.165 | -0.13 | -0.22, -0.04 | **0.007** | -0.11 | -0.21, -0.01 | 0.039 |
| Latinx | -0.03 | -0.12, 0.07 | 0.578 | -0.01 | -0.10, 0.08 | 0.844 | 0.01 | -0.08, 0.09 | 0.891 | 0.05 | -0.05, 0.16 | 0.324 | 0.04 | -0.07, 0.15 | 0.494 | -0.01 | -0.11, 0.08 | 0.766 | -0.05 | -0.11, 0.00 | 0.045 |
| Latinx-White | 0.10 | 0.00, 0.19 | 0.050 | 0.09 | -0.01, 0.19 | 0.092 | -0.03 | -0.12, 0.06 | 0.518 | 0.03 | -0.07, 0.12 | 0.599 | 0.01 | -0.10, 0.11 | 0.863 | -0.03 | -0.11, 0.04 | 0.417 | -0.01 | -0.08, 0.06 | 0.763 |
| Black+ | 0.00 | -0.09, 0.08 | 0.988 | 0.07 | -0.02, 0.15 | 0.111 | 0.15 | 0.06, 0.23 | **0.001** | 0.08 | -0.01, 0.18 | 0.087 | 0.07 | -0.02, 0.15 | 0.125 | 0.05 | -0.04, 0.14 | 0.240 | -0.01 | -0.07, 0.05 | 0.742 |
| Asian+ | -0.04 | -0.12, 0.03 | 0.284 | -0.06 | -0.14, 0.02 | 0.174 | -0.03 | -0.12, 0.06 | 0.499 | -0.01 | -0.10, 0.09 | 0.913 | -0.08 | -0.16, 0.00 | 0.038 | -0.03 | -0.11, 0.05 | 0.435 | 0.00 | -0.06, 0.06 | 0.983 |
|  | Controlling for family-level disadvantage |  |  |  |  |  |  |  |  |  |  |  |  |  |  |  |  |  |  |  |  |
|  | *b* | CI | *p* | *b* | CI | *p* | *b* | CI | *p* | *b* | CI | *p* | *b* | CI | *p* | *b* | CI | *p* | *b* | CI | *p* |
| Family-level disadvantage | 0.10 | -0.02, 0.23 | 0.106 | 0.18 | 0.06, 0.29 | **0.003** | 0.21 | 0.08, 0.33 | **0.001** | 0.02 | -0.10, 0.15 | 0.022 | 0.11 | 0.24, -0.03 | 0.122 | 0.08 | -0.04, 0.21 | 0.187 | 0.03 | -0.05, 0.10 | 0.508 |
| Latinx | -0.06 | -0.17, 0.05 | 0.286 | -0.06 | -0.17, 0.05 | 0.253 | -0.02 | -0.12, 0.09 | 0.762 | 0.01 | -0.11, 0.13 | 0.740 | 0.04 | 0.18, -0.09 | 0.538 | -0.02 | -0.12, 0.09 | 0.791 | -0.08 | -0.13, -0.03 | **0.001** |
| Latinx-White | 0.10 | -0.01, 0.21 | 0.077 | 0.07 | -0.06, 0.19 | 0.302 | -0.11 | -0.22, 0.00 | 0.055 | -0.03 | -0.15, 0.10 | 0.895 | -0.03 | 0.10, -0.16 | 0.684 | -0.03 | -0.10, 0.04 | 0.480 | 0.02 | -0.07, 0.11 | 0.726 |
| Black+ | -0.07 | -0.19, 0.05 | 0.264 | -0.02 | -0.12, 0.08 | 0.732 | 0.06 | -0.05, 0.16 | 0.311 | -0.06 | -0.18, 0.06 | 0.658 | 0.02 | 0.13, -0.10 | 0.775 | -0.03 | -0.14, 0.08 | 0.616 | -0.05 | -0.12, 0.02 | 0.172 |
| Asian+ | -0.04 | -0.13, 0.05 | 0.387 | -0.07 | -0.16, 0.02 | 0.138 | -0.08 | -0.17, 0.02 | 0.122 | 0.16 | 0.02, 0.29 | 0.315 | -0.10 | 0.00, -0.18 | 0.040 | -0.06 | -0.14, 0.02 | 0.129 | -0.04 | -0.08, 0.01 | 0.096 |

Standardized regression coefficients (*b*) and 95% confidence intervals (CIs) and uncorrected *p*-value calculated by regressing parent-reported mental health on socioeconomic inequality and racial/ethnic identity (with and without controlling for family-level disadvantage). *P*-values, where FDR corrected *p*-values < 0.05, are marked in bold. White-only identity is reference group. <sup>a</sup>All models included covariate adjustment for age, gender, and an age by gender interaction. *Note:* ADHD=attention-deficit hyperactivity disorder; ODD=oppositional defiant disorder; CD= conduct disorder.

**Table S4.** Associations between racial/ethnic identity and socioeconomic inequality.

|  | Family-level disadvantage |  |  | Neighborhood-level disadvantage |  |  | Neighborhood opportunity |  |  |
| --- | --- | --- | --- | --- | --- | --- | --- | --- | --- |
|  | *b* | 95% CI | *p* | *b* | 95% CI | *p* | *b* | 95% CI | *p* |
| Latinx | 0.25 | 0.16 – 0.35 | **<0.001** | 0.43 | 0.33– 0.52 | **<0.001** | -0.29 | -0.41– -0.17 | **<0.001** |
| Latinx-White | 0.02 | -0.09 – 0.12 | 0.763 | 0.04 | -0.06– 0.15 | 0.444 | 0.03 | -0.11– 0.16 | 0.700 |
| Black+ | 0.43 | 0.35 – 0.52 | **<0.001** | 0.33 | 0.25– 0.41 | **<0.001** | -0.28 | -0.38– -0.18 | **<0.001** |
| Asian+ | -0.10 | -0.19– -0.02 | **0.017** | -0.06 | -0.13– -0.01 | **0.037** | 0.09 | 0.01– 0.17 | 0.040 |

Standardized regression coefficients (*b*), 95% confidence intervals (CIs), and uncorrected *p*-values calculated by regressing socioeconomic measures on racial/ethnic identity. White-only identity is reference group. *P*-values, where FDR corrected *p*-values < 0.05, are marked in bold.

**Table S5.** Associations between socioeconomic inequality and racial/ethnic identity with DNA-methylation profiles.

|  | DNAm-CRP |  |  | DunedinPoAm |  |  |
| --- | --- | --- | --- | --- | --- | --- |
|  | No further covariates <sup>a</sup> |  |  |  |  |  |
|  | *b* | 95% CI | *p* | *b* | 95% CI | *p* |
| Family-level disadvantage | 0.22 | 0.12– 0.31 | **<0.001** | 0.28 | 0.18– 0.37 | **<0.001** |
| Neighborhood-level disadvantage | 0.25 | 0.17 – 0.33 | **<0.001** | 0.24 | 0.15 – 0.34 | **<0.001** |
| Neighborhood opportunity | -0.12 | -0.21– -0.03 | **0.012** | -0.19 | -0.30 – -0.08 | **0.001** |
| Latinx | 0.15 | 0.07– 0.24 | **0.001** | 0.18 | 0.10– 0.27 | **<0.001** |
| Latinx-White | 0.10 | -0.02– 0.22 | 0.108 | 0.09 | -0.01– 0.19 | 0.089 |
| Black+ | 0.08 | 0.02 – 0.15 | **0.012** | 0.19 | 0.11– 0.28 | **<0.001** |
| Asian+ | -0.01 | -0.09– 0.07 | 0.776 | 0.11 | -0.01– 0.22 | 0.058 |
|  | Controlling for BMI |  |  |  |  |  |
|  | *b* | 95% CI | *p* | *b* | 95% CI | *p* |
| BMI | 0.33 | 0.24– 0.42 | **<0.001** | 0.30 | 0.21– 0.40 | **<0.001** |
| Family-level disadvantage | 0.11 | 0.01– 0.21 | 0.024 | 0.19 | 0.08 – 0.29 | **<0.001** |
| Neighborhood-level disadvantage | 0.15 | 0.07– 0.23 | **<0.001** | 0.14 | 0.05 – 0.24 | **0.004** |
| Neighborhood opportunity | -0.05 | -0.15– 0.05 | 0.314 | -0.09 | -0.20 – 0.01 | 0.088 |
| Latinx | 0.10 | 0.01– 0.18 | 0.029 | 0.13 | 0.04– 0.22 | **0.004** |
| Latinx-White | 0.07 | -0.03– 0.17 | 0.180 | 0.06 | -0.03 – 0.14 | 0.213 |
| Black+ | 0.01 | -0.07– 0.07 | 0.975 | 0.11 | 0.02 – 0.19 | **0.011** |
| Asian+ | 0.01 | -0.07– 0.09 | 0.857 | 0.13 | 0.02 – 0.24 | **0.025** |
|  | Controlling for puberty |  |  |  |  |  |
|  | *b* | 95% CI | *p* | *b* | 95% CI | *p* |
| Puberty | 0.13 | 0.02– 0.24 | **0.020** | 0.05 | -0.07– 0.18 | 0.402 |
| Family-level disadvantage | 0.21 | 0.12 – 0.31 | **<0.001** | 0.28 | 0.18– 0.39 | **<0.001** |
| Neighborhood-level disadvantage | 0.25 | 0.16– 0.34 | **<0.001** | 0.25 | 0.11– 0.34 | **<0.001** |
| Neighborhood opportunity | -0.12 | -0.22 – -0.01 | **0.027** | -0.17 | -0.28– -0.06 | **0.003** |
| Latinx | 0.15 | 0.06– 0.24 | **0.001** | 0.18 | 0.09– 0.27 | **<0.001** |
| Latinx-White | 0.14 | 0.01 – 0.26 | **0.033** | 0.11 | 0.01– 0.21 | **0.035** |
| Black+ | 0.09 | 0.03 – 0.16 | **0.007** | 0.19 | 0.11– 0.27 | **<0.001** |
| Asian+ | -0.01 | -0.09 – 0.07 | 0.801 | 0.11 | -0.01 – 0.22 | 0.059 |
|  | Controlling for DNAm-smoke |  |  |  |  |  |
|  | *b* | 95% CI | *p* | *b* | 95% CI | *p* |
| DNAm-smoke | 0.40 | 0.28– 0.51 | **<0.001** | 0.24 | 0.11 – 0.37 | **<0.001** |
| Family-level disadvantage | 0.21 | 0.11– 0.30 | **<0.001** | 0.28 | 0.18– 0.38 | **<0.001** |
| Neighborhood-level disadvantage | 0.22 | 0.14– 0.30 | **<0.001** | 0.23 | 0.14 – 0.33 | **<0.001** |
| Neighborhood opportunity | -0.11 | -0.21– -0.02 | **0.022** | -0.17 | -0.27– -0.06 | **0.003** |
| Latinx | 0.11 | 0.03 – 0.20 | **0.011** | 0.16 | 0.07– 0.25 | **0.001** |
| Latinx-White | 0.12 | 0.01 – 0.23 | 0.048 | 0.11 | 0.01 – 0.20 | **0.028** |
| Black+ | 0.14 | 0.07 – 0.20 | **<0.001** | 0.22 | 0.14 – 0.31 | **<0.001** |
| Asian+ | -0.02 | -0.09 – 0.06 | 0.677 | 0.11 | -0.01 – 0.22 | 0.067 |
| Controlling for family-level disadvantage |  |  |  |  |  |  |
|  | *b* | 95% CI | *p* | *b* | 95% CI | *p* |
| Family-level disadvantage | 0.19 | 0.07– 0.30 | **0.002** | 0.24 | 0.13 – 0.36 | **<0.001** |
| Latinx | 0.10 | 0.01– 0.20 | 0.036 | 0.12 | 0.02 – 0.22 | **0.019** |
| Latinx-White | 0.12 | 0.00 – 0.23 | 0.050 | 0.10 | 0.01 – 0.20 | 0.047 |
| Black+ | 0.01 | -0.06 – 0.09 | 0.739 | 0.09 | -0.01 – 0.18 | 0.057 |
| Asian+ | 0.01 | -0.08 – 0.09 | 0.914 | 0.13 | 0.02 – 0.24 | **0.021** |

Standardized regression coefficients (*b*), 95% confidence intervals (CIs), and uncorrected p-values calculated by regressing DNA-methylation measures on socioeconomic measures and racial/ethnic identity with and without controlling for normed BMI z-scores, puberty (residualized for age within each gender), and DNA-methylation profiles of smoking (DNAm-smoke), separately. *P*-values, where FDR corrected *p*-values < 0.05, are marked in bold.
<sup>a</sup>All models included covariate adjustment for child's age, gender, and an age by gender interaction. Methylation scores were residualized for technical covariates (array, slide, batch, cell composition).

**Table S6.** Associations between socioeconomic inequality with DNA-methylation profiles for each racial/ethnic group.

|  | DNAm-CRP |  |  | DunedinPoAm |  |  |
| --- | --- | --- | --- | --- | --- | --- |
|  | *b* | 95% CI | White<br>*p* | *b* | 95% CI | *p* |
| Family-level disadvantage | 0.11 | -0.02– 0.24 | 0.098 | 0.23 | 0.08– 0.37 | 0.002 |
| Neighborhood-level disadvantage | 0.23 | 0.11– 0.35 | <0.001 | 0.21 | 0.06– 0.37 | 0.007 |
| Neighborhood opportunity | -0.07 | -0.19– 0.06 | 0.312 | -0.12 | -0.3– 0.06 | 0.182 |
|  | Latinx |  |  |  |  |  |
|  | *b* | 95% CI | *p* | *b* | 95% CI | *p* |
| Family-level disadvantage | 0.33 | 0.07– 0.59 | 0.014 | 0.118 | -0.18– 0.42 | 0.439 |
| Neighborhood-level disadvantage | 0.11 | -0.12– 0.33 | 0.352 | 0.01 | -0.27– 0.29 | 0.925 |
| Neighborhood opportunity | -0.18 | -0.43– 0.07 | 0.160 | -0.18 | -0.51– 0.15 | 0.278 |
|  | Latinx-White |  |  |  |  |  |
|  | *b* | 95% CI | *p* | *b* | 95% CI | *p* |
| Family-level disadvantage | 0.53 | 0.13– 0.93 | 0.010 | 0.29 | -0.07– 0.64 | 0.118 |
| Neighborhood-level disadvantage | 0.19 | -0.1– 0.47 | 0.197 | 0.11 | -0.16– 0.38 | 0.436 |
| Neighborhood opportunity | -0.01 | -0.20– 0.18 | 0.945 | -0.12 | -0.33– 0.08 | 0.232 |
|  | Black + |  |  |  |  |  |
|  | *b* | 95% CI | *p* | *b* | 95% CI | *p* |
| Family-level disadvantage | 0.06 | -0.22– 0.34 | 0.678 | 0.19 | -0.13– 0.51 | 0.252 |
| Neighborhood-level disadvantage | 0.49 | 0.22– 0.75 | <0.001 | 0.47 | 0.14– 0.79 | 0.005 |
| Neighborhood opportunity | -0.24 | -0.44– -0.04 | 0.018 | -0.25 | -0.52– 0.02 | 0.065 |
|  | Asian + |  |  |  |  |  |
|  | *b* | 95% CI | *p* | *b* | 95% CI | *p* |
| Family-level disadvantage | 0.08 | -0.18– 0.34 | 0.562 | 0.07 | -0.26– 0.41 | 0.668 |
| Neighborhood-level disadvantage | 0.07 | -0.2– 0.33 | 0.629 | -0.10 | -0.45– 0.25 | 0.590 |
| Neighborhood opportunity | 0.25 | -0.36– 0.86 | 0.428 | -0.02 | -0.85– 0.8 | 0.959 |

Standardized regression coefficients (*b*) and 95% confidence intervals (CIs) and uncorrected *p*-value calculated by regressing DNA-methylation measures on socioeconomic measures, separately, within each racial/ethnic group. All models included covariate adjustment for age, gender, and an age by gender interaction. Methylation scores were residualized for technical covariates (for methylation: array, slide, batch, cell composition).

**Table S7.** Associations between DNA-methylation and genetic profiles with parent-reported mental health.

|  | Internalizing |  |  | Attention Problems |  |  | Aggression |  |  | Rule-Breaking |  |  | ADHD |  |  | ODD |  |  | CD |  |  |
| --- | --- | --- | --- | --- | --- | --- | --- | --- | --- | --- | --- | --- | --- | --- | --- | --- | --- | --- | --- | --- | --- |
|  | *r* | CI | *p* | *r* | CI | *p* | *r* | CI | *p* | *r* | CI | *p* | *r* | CI | *p* | *r* | CI | *p* | *r* | CI | *p* |
| No further covariates<sup>a</sup> |  |  |  |  |  |  |  |  |  |  |  |  |  |  |  |  |  |  |  |  |  |
| DNA<sub>m</sub>-CRP | 0.15 | 0.05, 0.25 | **0.004** | 0.10 | -0.01, 0.21 | 0.083 | 0.17 | 0.04, 0.31 | **0.013** | 0.06 | -0.05, 0.16 | 0.270 | 0.09 | -0.01, 0.19 | 0.084 | 0.08 | -0.04, 0.19 | 0.194 | 0.16 | 0.02, 0.29 | 0.026 |
| DunedinPoAm | 0.15 | 0.05, 0.25 | **0.002** | 0.10 | 0.00, 0.20 | 0.058 | 0.11 | -0.02, 0.25 | 0.094 | 0.00 | -0.07, 0.08 | 0.948 | 0.11 | 0.00, 0.21 | 0.042 | 0.05 | -0.08, 0.17 | 0.459 | 0.05 | -0.09, 0.46 | 0.301 |
| PGS-CRP | 0.14 | 0.04, 0.23 | **0.004** | 0.16 | 0.05, 0.28 | **0.004** | 0.11 | -0.03, 0.23 | 0.115 | 0.09 | 0.00, 0.17 | 0.059 | 0.02 | -0.09, 0.12 | 0.766 | 0.05 | -0.06, 0.15 | 0.386 | 0.01 | -0.18, 0.19 | 0.953 |
| Controlling for BMI |  |  |  |  |  |  |  |  |  |  |  |  |  |  |  |  |  |  |  |  |  |
|  | *r* | CI | *p* | *r* | CI | *p* | *r* | CI | *p* | *r* | CI | *p* | *r* | CI | *p* | *r* | CI | *p* | *r* | CI | *p* |
| BMI | 0.13 | 0.04, 0.22 | **0.004** | -0.02 | -0.12, 0.07 | 0.669 | 0.14 | 0.02, 0.26 | 0.020 | 0.05 | -0.03, 0.14 | 0.201 | 0.09 | -0.01, 0.2 | 0.076 | 0.03 | -0.08, 0.15 | 0.589 | 0.09 | -0.1, 0.29 | 0.342 |
| DNA<sub>m</sub>-CRP | 0.08 | -0.03, 0.19 | 0.158 | 0.10 | -0.02, 0.23 | 0.112 | 0.10 | -0.05, 0.25 | 0.182 | 0.03 | -0.09, 0.15 | 0.627 | 0.04 | -0.08, 0.16 | 0.471 | 0.06 | -0.08, 0.19 | 0.405 | 0.11 | -0.02, 0.23 | 0.110 |
| DunedinPoAm | 0.10 | -0.01, 0.2 | 0.069 | 0.10 | -0.01, 0.22 | 0.085 | 0.05 | -0.1, 0.19 | 0.526 | -0.03 | -0.13, 0.07 | 0.583 | 0.08 | -0.04, 0.19 | 0.179 | 0.03 | -0.1, 0.15 | 0.668 | 0.15 | -0.1, 0.4 | 0.240 |
| PGS-CRP | 0.12 | 0.03, 0.22 | **0.009** | 0.17 | 0.06, 0.28 | **0.003** | 0.09 | -0.04, 0.22 | 0.182 | 0.08 | -0.01, 0.17 | 0.075 | 0.01 | -0.1, 0.11 | 0.934 | 0.05 | -0.06, 0.15 | 0.383 | 0.00 | -0.19, 0.18 | 0.974 |
| Controlling for pubertal status |  |  |  |  |  |  |  |  |  |  |  |  |  |  |  |  |  |  |  |  |  |
|  | *r* | CI | *p* | *r* | CI | *p* | *r* | CI | *p* | *r* | CI | *p* | *r* | CI | *p* | *r* | CI | *p* | *r* | CI | *p* |
| Puberty | -0.04 | -0.14, 0.06 | 0.435 | -0.14 | -0.25, -0.03 | **0.013** | -0.01 | -0.15, 0.12 | 0.865 | -0.03 | -0.13, 0.08 | 0.598 | -0.03 | -0.13, 0.07 | 0.568 | -0.04 | -0.17, 0.08 | 0.496 | -0.16 | -0.43, 0.11 | 0.245 |
| DNA<sub>m</sub>-CRP | 0.16 | 0.05, 0.26 | **0.004** | 0.12 | 0.01, 0.24 | 0.034 | 0.18 | 0.03, 0.32 | 0.016 | 0.06 | -0.07, 0.19 | 0.356 | 0.10 | -0.01, 0.2 | 0.076 | 0.08 | -0.04, 0.2 | 0.175 | 0.19 | 0.02, 0.35 | 0.025 |
| DunedinPoAm | 0.15 | 0.05, 0.25 | **0.002** | 0.11 | 0.01, 0.21 | 0.040 | 0.11 | -0.02, 0.25 | 0.098 | 0.01 | -0.09, 0.1 | 0.920 | 0.11 | 0.01, 0.22 | 0.041 | 0.05 | -0.07, 0.17 | 0.439 | 0.20 | -0.09, 0.48 | 0.180 |
| PGS-CRP | 0.14 | 0.04, 0.23 | **0.004** | 0.17 | 0.06, 0.28 | **0.003** | 0.10 | -0.03, 0.23 | 0.116 | 0.09 | 0, 0.18 | 0.051 | 0.02 | -0.09, 0.12 | 0.775 | 0.05 | -0.06, 0.15 | 0.374 | 0.01 | -0.18, 0.19 | 0.949 |
| Controlling for DNAm-smoke |  |  |  |  |  |  |  |  |  |  |  |  |  |  |  |  |  |  |  |  |  |
|  | *r* | CI | *p* | *r* | CI | *p* | *r* | CI | *p* | *r* | CI | *p* | *r* | CI | *p* | *r* | CI | *p* | *r* | CI | *p* |
| DNAm-smoke | -0.07 | -0.2, 0.06 | 0.301 | 0.05 | -0.09, 0.19 | 0.500 | -0.10 | -0.26, 0.06 | 0.217 | -0.03 | -0.15, 0.1 | 0.674 | -0.05 | -0.18, 0.08 | 0.454 | 0.06 | -0.07, 0.2 | 0.354 | -0.02 | -0.2, 0.17 | 0.850 |
| DNA<sub>m</sub>-CRP | 0.19 | 0.06, 0.31 | **0.004** | 0.08 | -0.07, 0.22 | 0.299 | 0.11 | -0.15, 0.37 | 0.400 | 0.08 | -0.05, 0.21 | 0.245 | 0.12 | -0.01, 0.24 | 0.069 | 0.05 | -0.09, 0.18 | 0.517 | 0.16 | 0.03, 0.3 | 0.021 |
| DunedinPoAm | 0.16 | 0.06, 0.26 | **0.002** | 0.08 | -0.02, 0.19 | 0.127 | 0.13 | -0.01, 0.27 | 0.076 | 0.00 | -0.09, 0.1 | 0.926 | 0.12 | 0.01, 0.23 | 0.031 | 0.03 | -0.1, 0.15 | 0.665 | 0.19 | -0.08, 0.45 | 0.177 |
| PGS-CRP | 0.14 | 0.04,<br>0.23 | **0.004** | 0.16 | 0.05,<br>0.27 | **0.005** | 0.11 | -0.02,<br>0.24 | 0.100 | 0.09 | 0,<br>0.11 | 0.057 | 0.02 | -0.09,<br>0.12 | 0.753 | 0.04 | -0.06,<br>0.15 | 0.411 | 0.01 | -0.18,<br>0.19 | 0.958 |
| Controlling for Family-Level Disadvantage |  |  |  |  |  |  |  |  |  |  |  |  |  |  |  |  |  |  |  |  |  |
|  | *r* | CI | *p* | *r* | CI | *p* | *r* | CI | *p* | *r* | CI | *p* | *r* | CI | *p* | *r* | CI | *p* | *r* | CI | *p* |
| Family-level disadvantage | 0.21 | 0.13,<br>0.3 | **0.000** | 0.14 | 0.04,<br>0.23 | **0.005** | 0.21 | 0.1,<br>0.32 | **0.000** | 0.10 | 0.02,<br>0.18 | 0.017 | 0.15 | 0.06,<br>0.25 | **0.001** | 0.13 | 0.04,<br>0.22 | **0.007** | 0.13 | -0.04,<br>0.29 | 0.136 |
| DNA-m-CRP | 0.09 | -0.01,<br>0.19 | 0.078 | 0.06 | -0.05,<br>0.17 | 0.301 | 0.12 | -0.03,<br>0.26 | 0.111 | 0.03 | -0.08,<br>0.14 | 0.593 | 0.05 | -0.06,<br>0.15 | 0.375 | 0.04 | -0.07,<br>0.15 | 0.472 | 0.12 | 0,<br>0.25 | 0.056 |
| DunedinPoAm | 0.08 | -0.02,<br>0.18 | 0.108 | 0.06 | -0.05,<br>0.16 | 0.289 | 0.05 | -0.09,<br>0.18 | 0.497 | -0.03 | -0.13,<br>0.06 | 0.490 | 0.06 | -0.05,<br>0.17 | 0.258 | 0.01 | -0.11,<br>0.12 | 0.935 | 0.15 | -0.11,<br>0.41 | 0.251 |
| PGS-CRP | 0.13 | 0.04,<br>0.21 | **0.005** | 0.16 | 0.05,<br>0.27 | **0.004** | 0.10 | -0.03,<br>0.22 | 0.135 | 0.08 | 0,<br>0.17 | 0.062 | 0.01 | -0.09,<br>0.12 | 0.813 | 0.04 | -0.06,<br>0.15 | 0.406 | 0.00 | -0.18,<br>0.19 | 0.962 |

Standardized regression coefficients (*r*) and 95% confidence intervals (CIs) and uncorrected *p*-value calculated by regressing parent-reported mental health on DNA-methylation measures and PGS-CRP with and without controlling for normed BMI z-scores, puberty (residualized for age within each gender), DNA-methylation profiles of smoking (DNA-m-smoke), and family-level disadvantage separately. *P*-values, where FDR corrected *p*-values < 0.05, are marked in bold. PGS analyses were restricted to participants of European ancestries as indicated by genetic ancestry PCs that are comparable to the GWAS discovery sample. <sup>a</sup>All models included covariate adjustment for age, gender, and an age by gender interaction. Methylation scores and PGS-CRP were residualized for technical covariates (for methylation: array, slide, batch, cell composition; for PGS-CRP: genetic ancestry PCs). *Note*: ADHD=attention-deficit hyperactivity disorder; ODD=oppositional defiant disorder; CD= conduct disorder.

**Table S8.**
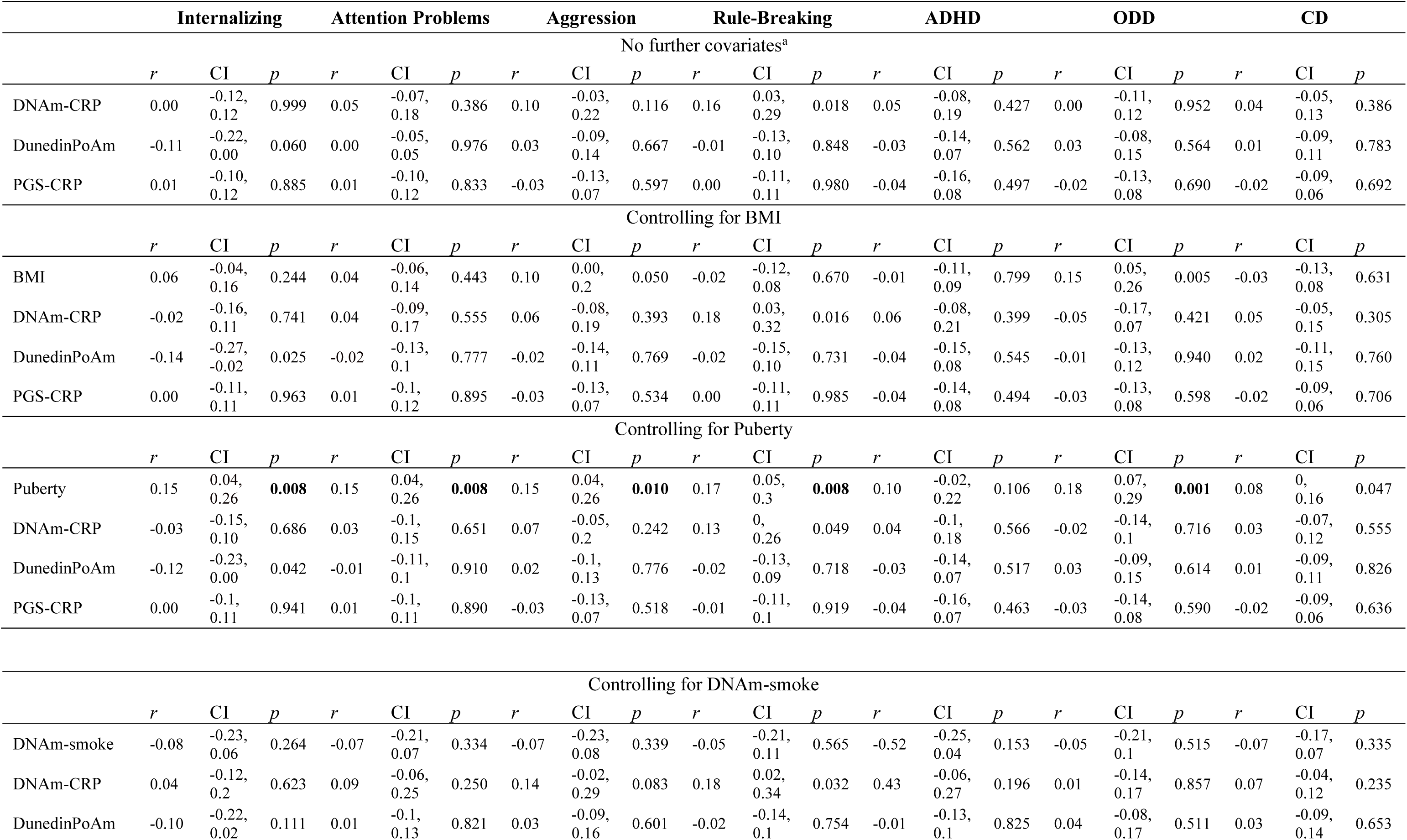
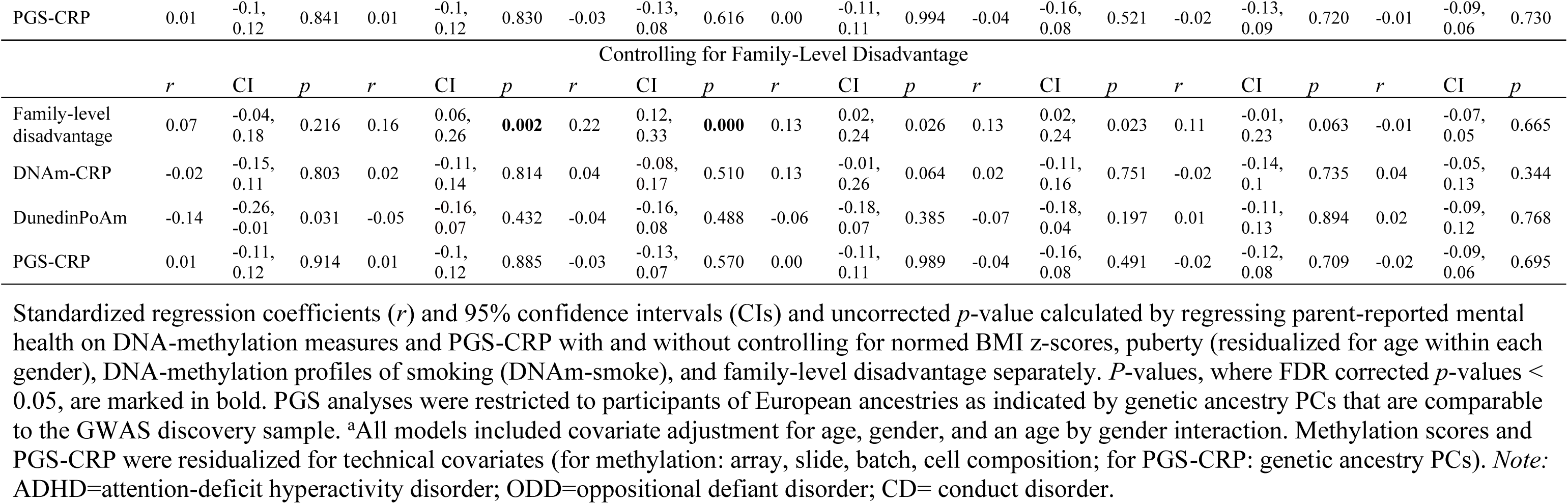
Associations between DNA-methylation and genetic profiles with self-reported mental health.

|  | Internalizing |  |  | Attention Problems |  |  | Aggression |  |  | Rule-Breaking |  |  | ADHD |  |  | ODD |  |  | CD |  |  |
| --- | --- | --- | --- | --- | --- | --- | --- | --- | --- | --- | --- | --- | --- | --- | --- | --- | --- | --- | --- | --- | --- |
| No further covariates<sup>a</sup> |  |  |  |  |  |  |  |  |  |  |  |  |  |  |  |  |  |  |  |  |  |
|  | *r* | CI | *p* | *r* | CI | *p* | *r* | CI | *p* | *r* | CI | *p* | *r* | CI | *p* | *r* | CI | *p* | *r* | CI | *p* |
| DNA<sub>m</sub>-CRP | 0.00 | -0.12, 0.12 | 0.999 | 0.05 | -0.07, 0.18 | 0.386 | 0.10 | -0.03, 0.22 | 0.116 | 0.16 | 0.03, 0.29 | 0.018 | 0.05 | -0.08, 0.19 | 0.427 | 0.00 | -0.11, 0.12 | 0.952 | 0.04 | -0.05, 0.13 | 0.386 |
| DunedinPoAm | -0.11 | -0.22, 0.00 | 0.060 | 0.00 | -0.05, 0.05 | 0.976 | 0.03 | -0.09, 0.14 | 0.667 | -0.01 | -0.13, 0.10 | 0.848 | -0.03 | -0.14, 0.07 | 0.562 | 0.03 | -0.08, 0.15 | 0.564 | 0.01 | -0.09, 0.11 | 0.783 |
| PGS-CRP | 0.01 | -0.10, 0.12 | 0.885 | 0.01 | -0.10, 0.12 | 0.833 | -0.03 | -0.13, 0.07 | 0.597 | 0.00 | -0.11, 0.11 | 0.980 | -0.04 | -0.16, 0.08 | 0.497 | -0.02 | -0.13, 0.08 | 0.690 | -0.02 | -0.09, 0.06 | 0.692 |
| Controlling for BMI |  |  |  |  |  |  |  |  |  |  |  |  |  |  |  |  |  |  |  |  |  |
|  | *r* | CI | *p* | *r* | CI | *p* | *r* | CI | *p* | *r* | CI | *p* | *r* | CI | *p* | *r* | CI | *p* | *r* | CI | *p* |
| BMI | 0.06 | -0.04, 0.16 | 0.244 | 0.04 | -0.06, 0.14 | 0.443 | 0.10 | 0.00, 0.2 | 0.050 | -0.02 | -0.12, 0.08 | 0.670 | -0.01 | -0.11, 0.09 | 0.799 | 0.15 | 0.05, 0.26 | 0.005 | -0.03 | -0.13, 0.08 | 0.631 |
| DNA<sub>m</sub>-CRP | -0.02 | -0.16, 0.11 | 0.741 | 0.04 | -0.09, 0.17 | 0.555 | 0.06 | -0.08, 0.19 | 0.393 | 0.18 | 0.03, 0.32 | 0.016 | 0.06 | -0.08, 0.21 | 0.399 | -0.05 | -0.17, 0.07 | 0.421 | 0.05 | -0.05, 0.15 | 0.305 |
| DunedinPoAm | -0.14 | -0.27, -0.02 | 0.025 | -0.02 | -0.13, 0.1 | 0.777 | -0.02 | -0.14, 0.11 | 0.769 | -0.02 | -0.15, 0.10 | 0.731 | -0.04 | -0.15, 0.08 | 0.545 | -0.01 | -0.13, 0.12 | 0.940 | 0.02 | -0.11, 0.15 | 0.760 |
| PGS-CRP | 0.00 | -0.11, 0.11 | 0.963 | 0.01 | -0.1, 0.12 | 0.895 | -0.03 | -0.13, 0.07 | 0.534 | 0.00 | -0.11, 0.11 | 0.985 | -0.04 | -0.14, 0.08 | 0.494 | -0.03 | -0.13, 0.08 | 0.598 | -0.02 | -0.09, 0.06 | 0.706 |
| Controlling for Puberty |  |  |  |  |  |  |  |  |  |  |  |  |  |  |  |  |  |  |  |  |  |
|  | *r* | CI | *p* | *r* | CI | *p* | *r* | CI | *p* | *r* | CI | *p* | *r* | CI | *p* | *r* | CI | *p* | *r* | CI | *p* |
| Puberty | 0.15 | 0.04, 0.26 | **0.008** | 0.15 | 0.04, 0.26 | **0.008** | 0.15 | 0.04, 0.26 | **0.010** | 0.17 | 0.05, 0.3 | **0.008** | 0.10 | -0.02, 0.22 | 0.106 | 0.18 | 0.07, 0.29 | **0.001** | 0.08 | 0, 0.16 | 0.047 |
| DNA<sub>m</sub>-CRP | -0.03 | -0.15, 0.10 | 0.686 | 0.03 | -0.1, 0.15 | 0.651 | 0.07 | -0.05, 0.2 | 0.242 | 0.13 | 0, 0.26 | 0.049 | 0.04 | -0.1, 0.18 | 0.566 | -0.02 | -0.14, 0.1 | 0.716 | 0.03 | -0.07, 0.12 | 0.555 |
| DunedinPoAm | -0.12 | -0.23, 0.00 | 0.042 | -0.01 | -0.11, 0.1 | 0.910 | 0.02 | -0.1, 0.13 | 0.776 | -0.02 | -0.13, 0.09 | 0.718 | -0.03 | -0.14, 0.07 | 0.517 | 0.03 | -0.09, 0.15 | 0.614 | 0.01 | -0.09, 0.11 | 0.826 |
| PGS-CRP | 0.00 | -0.1, 0.11 | 0.941 | 0.01 | -0.1, 0.11 | 0.890 | -0.03 | -0.13, 0.07 | 0.518 | -0.01 | -0.11, 0.1 | 0.919 | -0.04 | -0.16, 0.07 | 0.463 | -0.03 | -0.14, 0.08 | 0.590 | -0.02 | -0.09, 0.06 | 0.636 |
| Controlling for DNAm-smoke |  |  |  |  |  |  |  |  |  |  |  |  |  |  |  |  |  |  |  |  |  |
|  | *r* | CI | *p* | *r* | CI | *p* | *r* | CI | *p* | *r* | CI | *p* | *r* | CI | *p* | *r* | CI | *p* | *r* | CI | *p* |
| DNAm-smoke | -0.08 | -0.23, 0.06 | 0.264 | -0.07 | -0.21, 0.07 | 0.334 | -0.07 | -0.23, 0.08 | 0.339 | -0.05 | -0.21, 0.11 | 0.565 | -0.52 | -0.25, 0.04 | 0.153 | -0.05 | -0.21, 0.1 | 0.515 | -0.07 | -0.17, 0.07 | 0.335 |
| DNAm-CRP | 0.04 | -0.12, 0.2 | 0.623 | 0.09 | -0.06, 0.25 | 0.250 | 0.14 | -0.02, 0.29 | 0.083 | 0.18 | 0.02, 0.34 | 0.032 | 0.43 | -0.06, 0.27 | 0.196 | 0.01 | -0.14, 0.17 | 0.857 | 0.07 | -0.04, 0.12 | 0.235 |
| DunedinPoAm | -0.10 | -0.22, 0.02 | 0.111 | 0.01 | -0.1, 0.13 | 0.821 | 0.03 | -0.09, 0.16 | 0.601 | -0.02 | -0.14, 0.1 | 0.754 | -0.01 | -0.13, 0.1 | 0.825 | 0.04 | -0.08, 0.17 | 0.511 | 0.03 | -0.09, 0.14 | 0.653 |
| PGS-CRP | 0.01 | -0.1,<br>0.12 | 0.841 | 0.01 | -0.1,<br>0.12 | 0.830 | -0.03 | -0.13,<br>0.08 | 0.616 | 0.00 | -0.11,<br>0.11 | 0.994 | -0.04 | -0.16,<br>0.08 | 0.521 | -0.02 | -0.13,<br>0.09 | 0.720 | -0.01 | -0.09,<br>0.06 | 0.730 |
| Controlling for Family-Level Disadvantage |  |  |  |  |  |  |  |  |  |  |  |  |  |  |  |  |  |  |  |  |  |
|  | *r* | CI | *p* | *r* | CI | *p* | *r* | CI | *p* | *r* | CI | *p* | *r* | CI | *p* | *r* | CI | *p* | *r* | CI | *p* |
| Family-level disadvantage | 0.07 | -0.04,<br>0.18 | 0.216 | 0.16 | 0.06,<br>0.26 | **0.002** | 0.22 | 0.12,<br>0.33 | **0.000** | 0.13 | 0.02,<br>0.24 | 0.026 | 0.13 | 0.02,<br>0.24 | 0.023 | 0.11 | -0.01,<br>0.23 | 0.063 | -0.01 | -0.07,<br>0.05 | 0.665 |
| DNA-m-CRP | -0.02 | -0.15,<br>0.11 | 0.803 | 0.02 | -0.11,<br>0.14 | 0.814 | 0.04 | -0.08,<br>0.17 | 0.510 | 0.13 | -0.01,<br>0.26 | 0.064 | 0.02 | -0.11,<br>0.16 | 0.751 | -0.02 | -0.14,<br>0.1 | 0.735 | 0.04 | -0.05,<br>0.13 | 0.344 |
| DunedinPoAm | -0.14 | -0.26,<br>-0.01 | 0.031 | -0.05 | -0.16,<br>0.07 | 0.432 | -0.04 | -0.16,<br>0.08 | 0.488 | -0.06 | -0.18,<br>0.07 | 0.385 | -0.07 | -0.18,<br>0.04 | 0.197 | 0.01 | -0.11,<br>0.13 | 0.894 | 0.02 | -0.09,<br>0.12 | 0.768 |
| PGS-CRP | 0.01 | -0.11,<br>0.12 | 0.914 | 0.01 | -0.1,<br>0.12 | 0.885 | -0.03 | -0.13,<br>0.07 | 0.570 | 0.00 | -0.11,<br>0.11 | 0.989 | -0.04 | -0.16,<br>0.08 | 0.491 | -0.02 | -0.12,<br>0.08 | 0.709 | -0.02 | -0.09,<br>0.06 | 0.695 |

Standardized regression coefficients (*r*) and 95% confidence intervals (CIs) and uncorrected *p*-value calculated by regressing parent-reported mental health on DNA-methylation measures and PGS-CRP with and without controlling for normed BMI z-scores, puberty (residualized for age within each gender), DNA-methylation profiles of smoking (DNA-m-smoke), and family-level disadvantage separately. *P*-values, where FDR corrected *p*-values < 0.05, are marked in bold. PGS analyses were restricted to participants of European ancestries as indicated by genetic ancestry PCs that are comparable to the GWAS discovery sample. <sup>a</sup>All models included covariate adjustment for age, gender, and an age by gender interaction. Methylation scores and PGS-CRP were residualized for technical covariates (for methylation: array, slide, batch, cell composition; for PGS-CRP: genetic ancestry PCs). *Note*: ADHD=attention-deficit hyperactivity disorder; ODD=oppositional defiant disorder; CD= conduct disorder.

**Table S9.** Associations between DNA-methylation with parent-reported mental health for each racial/ethnic group.

|  | Internalizing |  |  | Attention Problems |  |  | Aggression |  |  | Rule-Breaking |  |  | ADHD |  |  | ODD |  |  | CD |  |  |
| --- | --- | --- | --- | --- | --- | --- | --- | --- | --- | --- | --- | --- | --- | --- | --- | --- | --- | --- | --- | --- | --- |
|  | White |  |  |  |  |  |  |  |  |  |  |  |  |  |  |  |  |  |  |  |  |
|  | *r* | CI | *p* | *r* | CI | *p* | *r* | CI | *p* | *r* | CI | *p* | *r* | CI | *p* | *r* | CI | *p* | *r* | CI | *p* |
| DNA<sub>m</sub>-CRP | 0.22 | 0.09, 0.34 | **0.001** | 0.08 | -0.06, 0.22 | 0.235 | 0.28 | 0.09, 0.46 | **0.003** | 0.09 | -0.05, 0.22 | 0.211 | 0.15 | 0.01, 0.29 | 0.038 | 0.10 | -0.04, 0.24 | 0.156 | 0.26 | -0.87, 1.39 | 0.652 |
| DunedinPoAm | 0.18 | 0.04, 0.31 | **0.010** | 0.12 | -0.02, 0.27 | 0.090 | 0.10 | -0.1, 0.3 | 0.320 | 0.06 | -0.07, 0.19 | 0.372 | 0.11 | -0.04, 0.25 | 0.157 | 0.01 | -0.14, 0.15 | 0.945 | -0.08 | -1.07, 0.9 | 0.869 |
|  | Latinx |  |  |  |  |  |  |  |  |  |  |  |  |  |  |  |  |  |  |  |  |
|  | *r* | CI | *p* | *r* | CI | *p* | *r* | CI | *p* | *r* | CI | *p* | *r* | CI | *p* | *r* | CI | *p* | *r* | CI | *p* |
| DNA<sub>m</sub>-CRP | 0.21 | -0.02, 0.44 | 0.069 | -0.03 | -0.37, 0.3 | 0.841 | 0.26 | -0.05, 0.58 | 0.104 | 0.08 | -0.33, 0.49 | 0.700 | 0.10 | -0.11, 0.31 | 0.339 | 0.03 | -0.15, 0.21 | 0.737 | -0.08 | -0.26, 0.11 | 0.405 |
| DunedinPoAm | 0.28 | 0.07, 0.49 | 0.009 | 0.00 | -0.34, 0.34 | 0.980 | 0.47 | 0.09, 0.84 | 0.014 | -0.02 | -0.26, 0.21 | 0.853 | 0.21 | 0.02, 0.4 | 0.034 | 0.02 | -0.12, 0.17 | 0.742 | 0.06 | -0.13, 0.26 | 0.521 |
|  | Latinx-White |  |  |  |  |  |  |  |  |  |  |  |  |  |  |  |  |  |  |  |  |
|  | *r* | CI | *p* | *r* | CI | *p* | *r* | CI | *p* | *r* | CI | *p* | *r* | CI | *p* | *r* | CI | *p* | *r* | CI | *p* |
| DNA<sub>m</sub>-CRP | -0.27 | -0.5, -0.04 | 0.020 | -0.01 | -0.32, 0.3 | 0.935 | -0.18 | -0.5, 0.13 | 0.252 | 0.07 | -0.14, 0.28 | 0.508 | -0.02 | -0.36, 0.33 | 0.917 | 0.19 | -0.24, 0.61 | 0.387 | 0.10 | -0.16, 0.36 | 0.438 |
| DunedinPoAm | -0.07 | -0.29, 0.15 | 0.523 | 0.05 | -0.26, 0.36 | 0.756 | -0.07 | -0.4, 0.26 | 0.681 | -0.11 | -0.35, 0.13 | 0.356 | 0.09 | -0.18, 0.35 | 0.525 | 0.12 | -0.18, 0.42 | 0.442 | 0.04 | -0.17, 0.25 | 0.693 |
|  | Black + |  |  |  |  |  |  |  |  |  |  |  |  |  |  |  |  |  |  |  |  |
|  | *r* | CI | *p* | *r* | CI | *p* | *r* | CI | *p* | *r* | CI | *p* | *r* | CI | *p* | *r* | CI | *p* | *r* | CI | *p* |
| DNA<sub>m</sub>-CRP | 0.17 | -0.44, 0.77 | 0.585 | 0.49 | -0.3, 1.27 | 0.223 | 0.12 | -0.5, 0.74 | 0.701 | -0.05 | -0.66, 0.56 | 0.872 | -0.21 | -0.62, 0.2 | 0.321 | -0.30 | -0.92, 0.32 | 0.342 | 0.52 | -0.23, 1.28 | 0.175 |
| DunedinPoAm | -0.04 | -0.4, 0.31 | 0.813 | -0.07 | -0.36, 0.22 | 0.645 | 0.04 | -0.39, 0.48 | 0.852 | -0.15 | -0.44, 0.15 | 0.326 | 0.14 | -0.23, 0.52 | 0.459 | 0.20 | -0.32, 0.73 | 0.450 | 0.68 | 0.13, 1.23 | 0.016 |
|  | Asian + |  |  |  |  |  |  |  |  |  |  |  |  |  |  |  |  |  |  |  |  |
|  | *r* | CI | *p* | *r* | CI | *p* | *r* | CI | *p* | *r* | CI | *p* | *r* | CI | *p* | *r* | CI | *p* | *r* | CI | *p* |
| DNA<sub>m</sub>-CRP | -0.14 | -0.42, 0.13 | 0.308 | -0.17 | -0.65, 0.31 | 0.484 | 0.04 | -0.27, 0.36 | 0.791 | -0.25 | -0.61, 0.12 | 0.186 | 0.04 | -0.16, 0.25 | 0.696 | -0.08 | -0.37, 0.22 | 0.614 | -0.11 | -0.38, 0.17 | 0.439 |
| DunedinPoAm | -0.28 | -0.63, 0.08 | 0.130 | -0.26 | -0.75, 0.23 | 0.294 | -0.29 | -0.67, 0.1 | 0.149 | -0.24 | -0.61, 0.13 | 0.200 | -0.08 | -0.34, 0.18 | 0.541 | -0.07 | -0.35, 0.22 | 0.648 | 0.55 | 0.3, 0.8 | **0.000** |

Standardized regression coefficients (*r*) and 95% confidence intervals (CIs) and uncorrected *p*-value calculated by regressing parent-reported mental health on DNA-methylation measures within each racial/ethnic group. <sup>a</sup>All models included covariate adjustment for age, gender, and an age by gender interaction. Methylation scores were residualized for technical covariates (array, slide, batch, cell composition). *Note:* ADHD=attention-deficit hyperactivity disorder; ODD=oppositional defiant disorder; CD= conduct disorder.

**Table S10.** Associations between DNA-methylation with self-reported mental health for each racial/ethnic group.

|  | Internalizing |  |  | Attention Problems |  |  | Aggression |  |  | Rule-Breaking |  |  | ADHD |  |  | ODD |  |  | CD |  |  |
| --- | --- | --- | --- | --- | --- | --- | --- | --- | --- | --- | --- | --- | --- | --- | --- | --- | --- | --- | --- | --- | --- |
| White |  |  |  |  |  |  |  |  |  |  |  |  |  |  |  |  |  |  |  |  |  |
|  | *r* | CI | *p* | *r* | CI | *p* | *r* | CI | *p* | *r* | CI | *p* | *r* | CI | *p* | *r* | CI | *p* | *r* | CI | *p* |
| DNA<sub>m</sub>-CRP | 0.07 | -0.1, 0.23 | 0.432 | 0.08 | -0.07, 0.23 | 0.278 | 0.073 | -0.08, 0.22 | 0.344 | 0.16 | 0.01, 0.32 | 0.038 | 0.12 | -0.04, 0.28 | 0.137 | 0.00 | -0.15, 0.14 | 0.956 | 0.08 | 0, 0.16 | 0.058 |
| DunedinPoAm | -0.12 | -0.29, 0.05 | 0.161 | 0.05 | -0.1, 0.19 | 0.522 | -0.01 | -0.16, 0.15 | 0.938 | 0.05 | -0.1, 0.2 | 0.497 | -0.01 | -0.16, 0.14 | 0.892 | 0.02 | -0.15, 0.2 | 0.795 | 0.10 | -0.01, 0.22 | 0.073 |
| Latinx |  |  |  |  |  |  |  |  |  |  |  |  |  |  |  |  |  |  |  |  |  |
|  | *r* | CI | *p* | *r* | CI | *p* | *r* | CI | *p* | *r* | CI | *p* | *r* | CI | *p* | *r* | CI | *p* | *r* | CI | *p* |
| DNA<sub>m</sub>-CRP | -0.40 | -0.76, -0.04 | 0.030 | -0.08 | -0.38, 0.22 | 0.587 | 0.05 | -0.37, 0.46 | 0.823 | -0.05 | -0.38, 0.29 | 0.790 | -0.24 | -0.66, 0.18 | 0.269 | -0.23 | -0.52, 0.06 | 0.115 | -0.14 | -0.9, 0.61 | 0.714 |
| DunedinPoAm | -0.03 | -0.29, 0.24 | 0.853 | 0.02 | -0.25, 0.29 | 0.903 | 0.14 | -0.11, 0.38 | 0.266 | -0.06 | -0.26, 0.14 | 0.548 | -0.07 | -0.33, 0.2 | 0.616 | 0.05 | -0.2, 0.3 | 0.705 | -0.40 | -1.11, 0.3 | 0.260 |
| Latinx-White |  |  |  |  |  |  |  |  |  |  |  |  |  |  |  |  |  |  |  |  |  |
|  | *r* | CI | *p* | *r* | CI | *p* | *r* | CI | *p* | *r* | CI | *p* | *r* | CI | *p* | *r* | CI | *p* | *r* | CI | *p* |
| DNA<sub>m</sub>-CRP | -0.30 | -0.67, 0.07 | 0.110 | 0.03 | -0.27, 0.33 | 0.845 | 0.07 | -0.34, 0.48 | 0.740 | -0.05 | -0.35, 0.26 | 0.768 | -0.14 | -0.56, 0.28 | 0.511 | 0.31 | -0.17, 0.78 | 0.207 | -0.41 | -0.83, 0.01 | 0.058 |
| DunedinPoAm | -0.02 | -0.28, 0.24 | 0.889 | 0.03 | -0.24, 0.3 | 0.839 | 0.14 | -0.11, 0.39 | 0.262 | -0.01 | -0.26, 0.14 | 0.533 | -0.03 | -0.31, 0.25 | 0.828 | 0.08 | -0.14, 0.3 | 0.463 | -0.22 | -0.94, 0.51 | 0.559 |
| Black + |  |  |  |  |  |  |  |  |  |  |  |  |  |  |  |  |  |  |  |  |  |
|  | *r* | CI | *p* | *r* | CI | *p* | *r* | CI | *p* | *r* | CI | *p* | *r* | CI | *p* | *r* | CI | *p* | *r* | CI | *p* |
| DNA<sub>m</sub>-CRP | 0.16 | -0.39, 0.7 | 0.575 | -0.28 | -0.95, 0.39 | 0.411 | 0.33 | -0.35, 1.01 | 0.344 | 0.29 | -0.3, 0.89 | 0.330 | -0.04 | -0.8, 0.71 | 0.917 | 0.27 | -0.16, 0.69 | 0.218 | 1.01 | -2.43, 4.46 | 0.564 |
| DunedinPoAm | -0.12 | -0.41, 0.16 | 0.393 | -0.28 | -0.64, 0.08 | 0.125 | 0.18 | -0.18, 0.53 | 0.325 | -0.07 | -0.35, 0.22 | 0.637 | -0.27 | -0.58, 0.04 | 0.089 | 0.28 | -0.06, 0.62 | 0.104 | -0.05 | -2.92, 2.82 | 0.972 |
| Asian + |  |  |  |  |  |  |  |  |  |  |  |  |  |  |  |  |  |  |  |  |  |
|  | *r* | CI | *p* | *r* | CI | *p* | *r* | CI | *p* | *r* | CI | *p* | *r* | CI | *p* | *r* | CI | *p* | *r* | CI | *p* |
| DNA<sub>m</sub>-CRP | 0.13 | -0.24, 0.49 | 0.496 | 0.14 | -0.24, 0.51 | 0.474 | 0.09 | -0.32, 0.49 | 0.664 | 0.08 | -0.33, 0.49 | 0.714 | -0.21 | -0.47, 0.04 | 0.103 | -0.19 | -0.53, 0.14 | 0.255 | -0.11 | -0.55, 0.33 | 0.619 |
| DunedinPoAm | -0.20 | -0.62, 0.22 | 0.355 | -0.10 | -0.56, 0.37 | 0.689 | -0.28 | -0.77, 0.2 | 0.252 | -0.25 | -0.66, 0.17 | 0.251 | -0.27 | -0.56, 0.03 | 0.073 | -0.40 | -0.72, -0.08 | 0.014 | -0.28 | -0.78, 0.22 | 0.269 |

Standardized regression coefficients (*r*) and 95% confidence intervals (CIs) and uncorrected *p*-value calculated by regressing self-reported mental health on DNA-methylation measures within each racial/ethnic group. <sup>a</sup>All models included covariate adjustment for age, gender, and an age by gender interaction. Methylation scores were residualized for technical covariates (array, slide, batch, cell composition). *Note:* ADHD=attention-deficit hyperactivity disorder; ODD=oppositional defiant disorder; CD= conduct disorder.

**Table S11.** Co-twin-control associations between DNA-methylation and parent-reported mental health.

|  | DNAm-CRP |  |  | DunedinPoAm |  |  |
| --- | --- | --- | --- | --- | --- | --- |
|  | *rA* | CI | *p* | *rA* | CI | *p* |
| Internalizing | 0.28 | -0.10, 0.66 | 0.146 | 0.30 | -0.19, 0.78 | 0.226 |
| Attention problems | 0.23 | -0.24, 0.70 | 0.339 | 0.21 | -0.38, 0.80 | 0.488 |
| Aggression | 0.30 | 0.02, 0.58 | **0.037** | 0.15 | -0.23, 0.53 | 0.431 |
| Rule-breaking | 0.19 | -0.03, 0.41 | 0.086 | 0.12 | -0.14, 0.38 | 0.375 |
| ADHD | 0.05 | -0.09, 0.18 | 0.505 | 0.077 | -0.08, 0.24 | 0.340 |
| ODD | 0.08 | -0.10, 0.26 | 0.370 | 0.03 | -0.30, 0.36 | 0.854 |
| CD | 0.26 | -0.50, 1.02 | 0.502 | 0.40 | -0.43, 1.22 | 0.348 |
|  | *rC* | CI | *p* | *rC* | CI | *p* |
| Internalizing | -1 | -1, -1 | <0.001 | -0.48 | -5.72, 4.75 | 0.857 |
| Attention problems | -1 | -1, -1 | <0.001 | 0.18 | -2.70, 3.06 | 0.904 |
| Aggression | 1 | 1, 1 | <0.001 | 1 | 1, 1 | <0.001 |
| Rule-breaking | -0.96 | -1.71, -0.22 | 0.011 | 1 | 1, 1 | <0.001 |
| ADHD | -0.96 | -0.96, -0.96 | <0.001 | 1 | 1, 1 | <0.001 |
| ODD | -0.78 | -3.34, 1.78 | 0.553 | 1 | 1, 1 | <0.001 |
| CD | -1 | -1, -1 | <0.001 | -1 | -1, -1 | <0.001 |
|  | *rE* | CI | *p* | *rE* | CI | *p* |
| Internalizing | -0.07 | -0.19, 0.05 | 0.248 | -0.03 | -0.16, 0.10 | 0.671 |
| Attention problems | -0.04 | -0.17, 0.10 | 0.591 | -0.05 | -0.18, 0.08 | 0.470 |
| Aggression | -0.12 | -0.27, 0.03 | 0.117 | -0.14 | -0.28, 0.01 | 0.062 |
| Rule-breaking | -0.09 | -0.22, 0.03 | 0.145 | -0.11 | -0.23, 0.01 | 0.075 |
| ADHD | 0.08 | -0.05, 0.21 | 0.221 | 0.09 | -0.02, 0.20 | 0.106 |
| ODD | 0.02 | -0.09, 0.12 | 0.766 | -0.04 | -0.13, 0.06 | 0.424 |
| CD | 0.04 | -0.07, 0.15 | 0.436 | 0.01 | -0.09, 0.10 | 0.915 |

Regression coefficients, 95% confidence intervals (CIs), and *p*-value calculated in a bivariate biometric model that decomposed the association between DNA-methylation and cognition into components representing additive genetic factors (*A*), environmental factors shared by twins living in the same home (*C*), and environmental factors unique to each twin (*E*). *rA* is the correlation between the *A* components of variation in DNA-methylation and cognition, which reflects the extent to which genetic variation in DNAm accounts for differences in cognitive functioning. *rC* is the correlation between the *C* components of variation in DNA-methylation and cognition, which reflects the extent to which shared environmental variation in DNAm accounts for differences in cognitive functioning. *rE* is the correlation between the *E* components of variation in DNA-methylation and cognition, which reflects the extent to which identical twins who differ from their co-twins in DNAm show corresponding differences in their cognitive functioning. *Note:* ADHD=attention-deficit hyperactivity disorder; ODD=oppositional defiant disorder; CD= conduct disorder.

## Notes

### Competing Interest Statement

The authors have declared no competing interest.

### Author Declarations

The University of Texas at Austin Institutional Review board granted ethical approval.

### Summary of Updates

Updated email address

